# Body mass index in parents and their adult offspring: a systematic review and meta-analysis

**DOI:** 10.1101/2022.08.01.22278184

**Authors:** Jie Zhang, Gemma Clayton, Kim Overvad, Anja Olsen, Deborah A Lawlor, Christina C Dahm

## Abstract

Obesity may track across generations, due to genetics and family environmental factors, or possibly intrauterine programming. However, many studies only assess associations between maternal body mass index (BMI) on offspring obesity in childhood. To determine whether maternal and paternal associations with offspring BMI differ, and whether associations persist into adulthood, a systematic review and meta-analysis was done. MEDLINE, EMBASE and PubMed (to December 2019) were searched. Observational studies reporting associations between maternal or paternal BMI and adult offspring BMI were included. Offspring BMIs were reported as continuous or categorical measures. 46 studies were included in the systematic review. Meta-analyses were conducted using random effects models. Parental BMI was positively associated with offspring BMI in adulthood. The pooled mother-offspring standardized mean difference (SMD) was 0.23 (95% confidence interval (CI): 0.20, 0.26), and father-offspring SMD was similar: 0.22 (95%CI: 0.19, 0.25) in adjusted models. Maternal and paternal overweight and obesity were associated with higher offspring BMI with similar magnitudes. If these associations are causal, they support interventions targeting all family members, rather than focusing solely on mothers, to maintain a healthy weight.

## 1. INTRODUCTION

Obesity represents one of the most urgent national and global health challenges because of its high prevalence and long-term adverse health consequences^1^. Familial resemblance in obesity is well-recognized^2^. To date, multiple studies have explored parental-offspring body mass index (BMI) associations and linked maternal or paternal adiposity with unfavorable offspring body composition and related morbidity (e.g. diabetes, cardiovascular and metabolic diseases)^3,4^.

However, the causes of the intergenerational association are unclear. Genetic predisposition, family socioeconomic status, and lifestyle factors (e.g., diet, physical activity) are important contributors to risk of offspring adiposity^5,6^. In addition to the above factors, mothers with greater BMI during pregnancy might increase the risk of obesity in the offspring via the fetal environment according to the ‘fetal overnutrition’ hypothesis^7,8^. The hypothesis suggests that maternal nutritional imbalance during gestation can alter metabolic processes, including the hypothalamic response to leptin and subsequent regulation of appetite and pancreatic beta-cell physiology, and have a persistent effect on offspring adiposity^9^. This hypothesis is supported by some animal studies^10,11^, yet the evidence is mixed in human studies^12,13^. However, novel evidence from Mendelian randomization studies which use maternal genetic variants adjusted for offspring genetic variants as unconfounded instrumental variables for an intrauterine exposure casts doubt on the causal effect of maternal pregnancy adiposity^14,15^. Furthermore, ‘negative control’ studies, which used paternal adiposity as a control and compared mother-offspring and father-offspring BMI associations suggest that maternal associations may be explained by family confounding^6,16^. If there is a causal *in-utero* effect, the association between parental-offspring BMI should be stronger in the maternal line than in the paternal line^17^. There is also growing interest to investigate whether any such association differs between daughters and sons. Comparison between the same-sex (mother-daughter, father-son) versus cross-sex (mother-son, father-daughter) transmission would provide more insight into understanding the mechanisms underlying the association. If a stronger same-sex association is observed, it might imply that shared environment rather than genes is the driver because selective mother-daughter and father-son gene transmission is not a common Mendelian trait^18^. However, no consistent patterns have been identified so far, making it difficult to draw conclusions. While previous systematic reviews have assessed associations between parental BMI and BMI of the offspring during childhood^6,19^, few have investigated sex-specific parental-offspring BMI associations, and very little is known about the strength of associations with offspring in adulthood. We undertook a systematic review and meta-analyses to quantify the strengths of associations between parental BMI and adult offspring obesity, including separately by maternal and paternal lines. The objectives of the present study were 3-fold: 1) to synthesize data from studies published from 1980 to 2020, thus investigating whether parental BMI is persistently associated with offspring adiposity in adulthood; 2) to explore the difference in maternal or paternal associations with adult offspring BMI and compare the associations for maternal and paternal lines; 3) to assess same-sex (mother-daughter, father-son) and cross-sex (mother-son, father-daughter) parent-offspring BMI associations.

## 2. METHODS

The meta-analysis was performed following the Meta-analysis of Observational Studies in Epidemiology (MOOSE) recommendation^20^. The study was registered in PROSPERO (CRD42020159281), an international database of prospectively registered systematic reviews.

### 2.1 Search strategy

We carried out a systematic literature search of PubMed, Embase, Web of Science, COCHRANE, and Google Scholar using predefined search terms (Appendix). Searches were restricted to human studies published in English, full-text articles published after 1980. The reference lists of all studies that met the inclusion criteria and all related systematic reviews were searched by J.Z., H.T.V, and C.C.D. Citation searches for all studies that met the inclusion criteria and all related systematic reviews were performed using Google Scholar Citations. Authors of included studies were contacted for additional data when required for inclusion in the meta-analyses. Database searches were completed in August 2019 and updated in July 2020.

### 2.2 Study eligibility criteria

#### 2.2.1 Inclusion criteria

Inclusion criteria were peer-reviewed studies reporting both the exposure variable (maternal BMI or paternal BMI, overweight, or obesity) and the outcome variable (offspring BMI, overweight, or obesity) among offspring aged 18 years or older. Exposure and outcome variables could be either continuous or categorical. To be included, studies had to have been published as full articles written in English, and express the findings as correlation or regression coefficients with 95% confidence intervals (CIs), or odds ratios (OR) or risk ratios (RR) with 95% CI. Studies reporting offspring BMI in late adolescence/early adulthood (reporting any offspring aged 18 years or older) were included. We included the first two generations, that is grandparents and parents, in a three-generational cohort study where the youngest generation was less than 18 years of age. Continuous BMI was selected as the main measurement of adiposity in meta-analyses. Where studies were based on the same cohort (e.g 1958 British birth cohort) and the same participants, we chose the study with the larger study population or older age group.

#### 2.2.2 Study selection

Three assessors (J.Z., H.T.V, M.L.L.) independently reviewed the titles and abstracts of all identified citations using Covidence online software (Covidence.org)^21^. Each full-text article was independently evaluated by three reviewers (J.Z., C.C.D, H.T.V). Disagreements were settled by discussion and consensus, with the third reviewer (C.C.D) available as an adjudicator.

#### 2.2.3 Data extraction

A piloted data collection form was used by three reviewers (J.Z., C.C.D., H.T.V.) to independently extract the following data from full-text articles: study design, years of study, study origin (country), study setting, sample size, assessment methods of weight and height information, when BMI was ascertained, whether maternal BMI pertained to pre-pregnancy, and reported measures of association. We included information available from the publications. Inconsistencies were checked and resolved through the consensus process described earlier.

### 2.3 Data synthesis

#### 2.3.1 Continuous outcome

We extracted correlation coefficients (r) or regression coefficients (mean difference (MD) or standardized mean difference (SMD)) for continuous outcomes. As studies varied in reporting associations between different familial groupings, we summarized three levels of family relationship groups: i) sex-specific at parental level and offspring level: mother-daughter, mother-son, father-daughter, and father-son; (ii) sex-specific at parental level: mother-offspring and father-offspring. When measures of association were reported by offspring sex at parental and offspring level, we used methods proposed by Borenstein et al.^22^ to combine father-offspring (or mother-offspring) from sex-specific associations (father-daughter and father-son, or mother-daughter and mother-son) and assumed varying correlations (ranging from 0-0.3) depending on the study design and analysis of the study to account for dependency. For example, we assumed a correlation of 0.3 where multilevel models had accounted for the structure of the data whilst we assumed a correlation of 0 in models with e.g. fathers with only one child. Inverse variance-weighted random-effects modelling was conducted to pool the sex-specific associations to either father-offspring or mother-offspring. Finally, random-effect meta-analysis was applied to synthesize the overall parental-offspring association since it accounts for both random variability and the variability in effects among the studies.

For data from regression models (where offspring BMI could be in kg/m^2^ or SD units), we separately extracted regression coefficients that were (i) MD per kg/m^2^ difference in parental BMI (ii) SMD per SD difference in parental BMI^23-26^. We then transformed studies reporting MD per kg/m^2^ difference in parental BMI to SD units, so that (i) and (ii) could be combined to increase power. Regression coefficients from multilevel models were treated as standard regression models. For studies reporting correlation coefficients (r), data were transformed with Fisher’s Z scale, and via the inverse Fisher’s Z transformation to r. The correlation coefficients were combined with unadjusted regression coefficients for the unadjusted data synthesis. The unadjusted and adjusted data (minimally adjusted for offspring’s age and sex) were synthesized separately.

Furthermore, we tested whether there were differences between mother-offspring and father-offspring associations. Differences in SMD or MD were calculated for continuous outcomes. Forest plots were constructed to display the individual and aggregate measures of association and their corresponding 95% CIs, in addition to tables presenting the aggregate results. We calculated the I^2^ value to measure heterogeneity, which represents the observed percentage of total variation across studies due to heterogeneity rather than chance.

#### 2.3.2 Categorical outcome

ORs and RRs and either their standard errors or 95%CI were directly extracted from study results. Because few studies reported obesity prevalence of more than 20% or reported RRs, ORs and RRs were summarized together^27^. Studies varied in reporting parental or offspring weight status using different criteria, different reference groups (e.g., normal weight mother, normal weight father, both normal weight parents), or varied in stratifying sex-specific associations (e.g. mother/father-daughter, mother/father-son association, parent-daughter/parent-son association, or mother/father-offspring, or parent-offspring), precluding synthesizing results to a meaningful single summary OR. We therefore presented forest plots showing individual study results without pooling ORs.

### 2.4 Subgroup analyses

The following subgroup analyses were specified a priori for the primary analysis, where data were available: (i) BMI assessment method: measured versus self-reported; (ii) study design: cross-sectional study versus longitudinal study; (iii) maternal BMI measured before pregnancy versus after pregnancy; (iv) offspring age: early adulthood (younger than 30 years), mid-adulthood (30-40years), and later adulthood (older than 40 years).

### 2.5 Quality assessment

Quality assessment was done by two reviewers (J.Z. and C.C.D) using a Newcastle-Ottawa scale (Appendix). The Newcastle-Ottawa scale evaluated the study design, representativeness of the exposed cohort, selection of the non-exposed cohort, ascertainment of exposure, comparability of cohorts on the basis of the design or analysis, assessment of offspring BMI, whether all adult participants in the analysis were the target age of the research question, and the adequacy of follow up of cohorts. We also considered how missing data were handled, whether studies included only biological parents and children (non-paternity), and adjustment for key confounders in the quality assessment. Studies were awarded one point per fulfilled criteria.

### 2.6 Post hoc analyses and publication bias

We conducted post hoc analyses by excluding 7 studies^26,28-33^ (15% of studies and 5% of participants) that measured offspring BMI at 15–25 years or took out 3 studies^25,29,34^ with low quality scores. To evaluate the potential effect of unpublished studies on our main findings (due to asymmetry or publication bias), we produced funnel plots and conducted Egger’s regression^35^.

All analyses were conducted using the metan command in Stata BE version 17^36^ (StataCorp, College Station, TX, USA).

## 3. RESULTS

### 3.1 Descriptive statistics

The searches identified 32,689 studies, of which 90 studies met the abstract inclusion criteria. Of the studies that met the initial abstract screening, 44 studies were excluded from the review, because: study subjects were not adult offspring (n=22), wrong exposure or outcome (n=10), adiposity was not assessed by BMI (n=3), exposure or outcome were BMI change (n=4), articles were not in English (n=1), wrong comparator (n=1), wrong study design (n=1), same cohort (n=1), or conference abstract (n=1). This resulted in 46 publications in the review (*Figure S1*). Of these studies, 26 were cohort studies, and 20 were cross-sectional studies. Study populations were from Australia, Asia, Europe, North and South America. Most of the studies included parent-offspring pairs and trios, with sample size ranging from 32 mother– daughter dyads^29^ to 36,528 father-mother-offspring trios^37^. Two studies used a three-generation cohort^38,39^. Parental and offspring BMIs were examined at different life stages across studies. Some studies focused on parental BMI at an early age^24,30,40-43^, while other studies collected parental anthropometric information in parallel with that of the adult offspring, so that parents were in middle or late adulthood ^23,25,31,32,44-46^. The information extracted from each study is presented in *Table S1*.

Various statistical methods were employed to analyze the relationship between parental BMI and adult offspring BMI. These included correlation coefficient methods, linear regression models, logistic or multinomial models, and multilevel models. Most linear regression studies adjusted for family socioeconomic factors, maternal age, smoking, drinking, and other important confounders. The majority of studies found positive associations between parents and their adult offspring BMI, while four studies reported null associations^25,34,38,46^. Several studies investigated sex-specific associations and compared the same-sex (mother-daughter, or father-son) or cross-sex (mother-son, father-daughter) relationships within the same population^18,23,24,47,48^. Conclusions regarding the findings for these sex-specific results varied between the studies. Detailed information is found in *Table S2, S3, and S4*.

### 3.2 Standardized Mean Difference (SMD) for continuous measures of BMI

Associations of continuous measures of maternal and/or paternal BMI with offspring BMI were reported in 35 studies. These studies used different BMI units and not all adjusted for confounders. We pooled results separately for: (i) confounder-adjusted SMD (n = 15 studies); (ii) unadjusted (for any covariates) SMD (n = 21) and (iii) confounder-adjusted associations with parental and offspring BMI in 1 kg/m^2^ units (n= 9).

Across all of these studies, results were consistent with similar positive associations for mother-offspring and father-offspring. For example, the pooled analyses showed mother-offspring SMD was 0.23 (95%CI: 0.20, 0.26 per SD greater maternal BMI), and the father-offspring was 0.22 (95%CI: 0.19, 0.25 per SD greater paternal BMI) in adjusted models (*Figure 1*). The equivalent unadjusted SMD results were 0.24 (95%CI: 0.21, 0.28) for mother-offspring and 0.21(95% CI: 0.18, 0.25) for father-offspring (*Figure S2*, with additional result in *Table S5*). There was evidence of between study heterogeneity in both analyses (I^2^% =79% for mother-offspring, I^2^%=68% for father-offspring). The synthesized sex-specific adjusted SMDs revealed similar positive associations for mother-daughter, mother-son, father-daughter, and father-son (*Figure 2, Table S5*).

**Figure 1.**
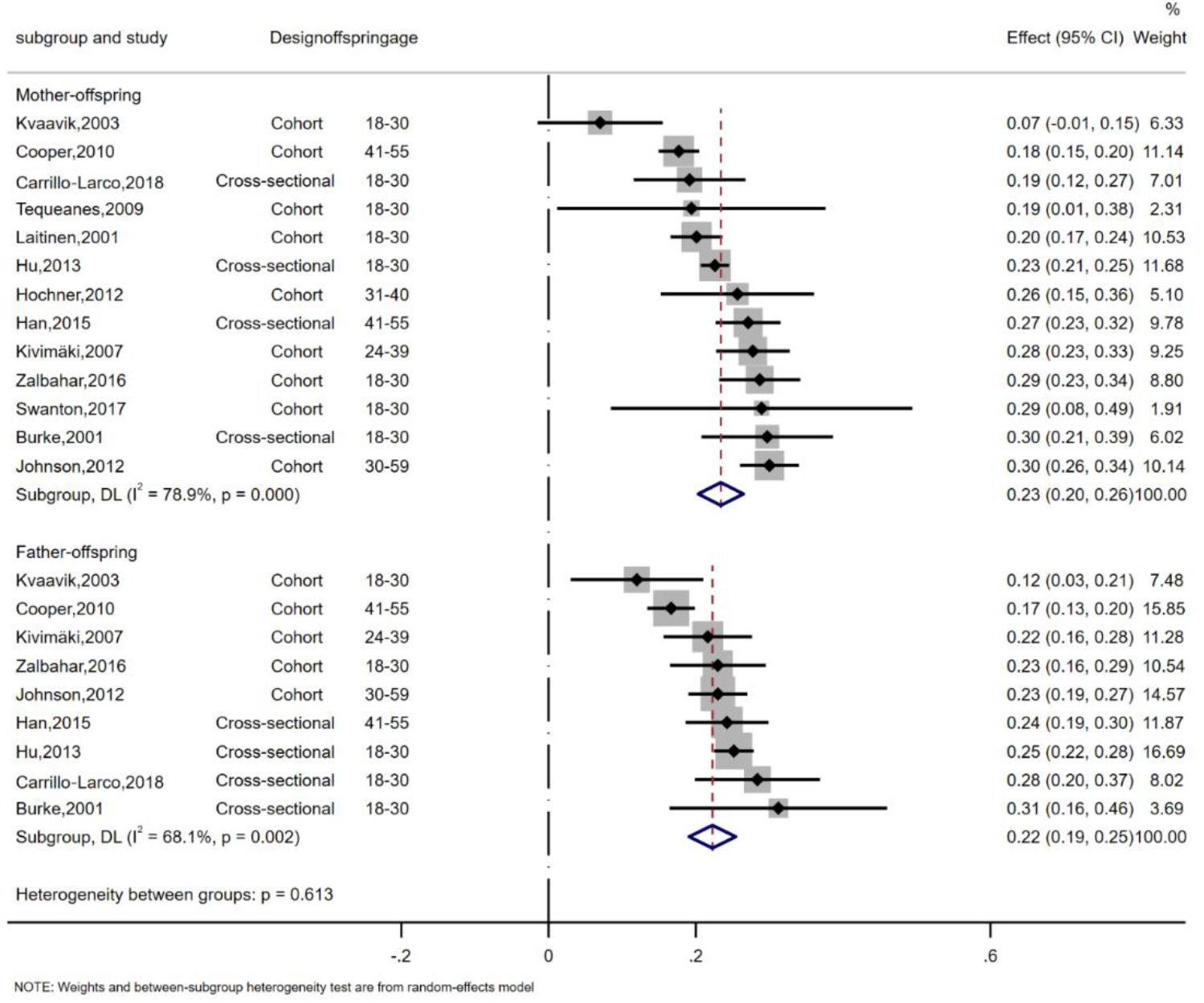
Meta-analysis of the association between parent and offspring BMI (standardized mean difference) The effect size estimate is the standardized mean difference with 95% confidence intervals (per SD of parental BMI) The size of the squares estimates is proportional to the weight assigned to each study. Diamonds represent pooled estimates from a random effects meta-analysis. The *I*^2^ and *P* values for heterogeneity are shown. BMI, body mass index; CI, confidence interval

**Figure 2.**
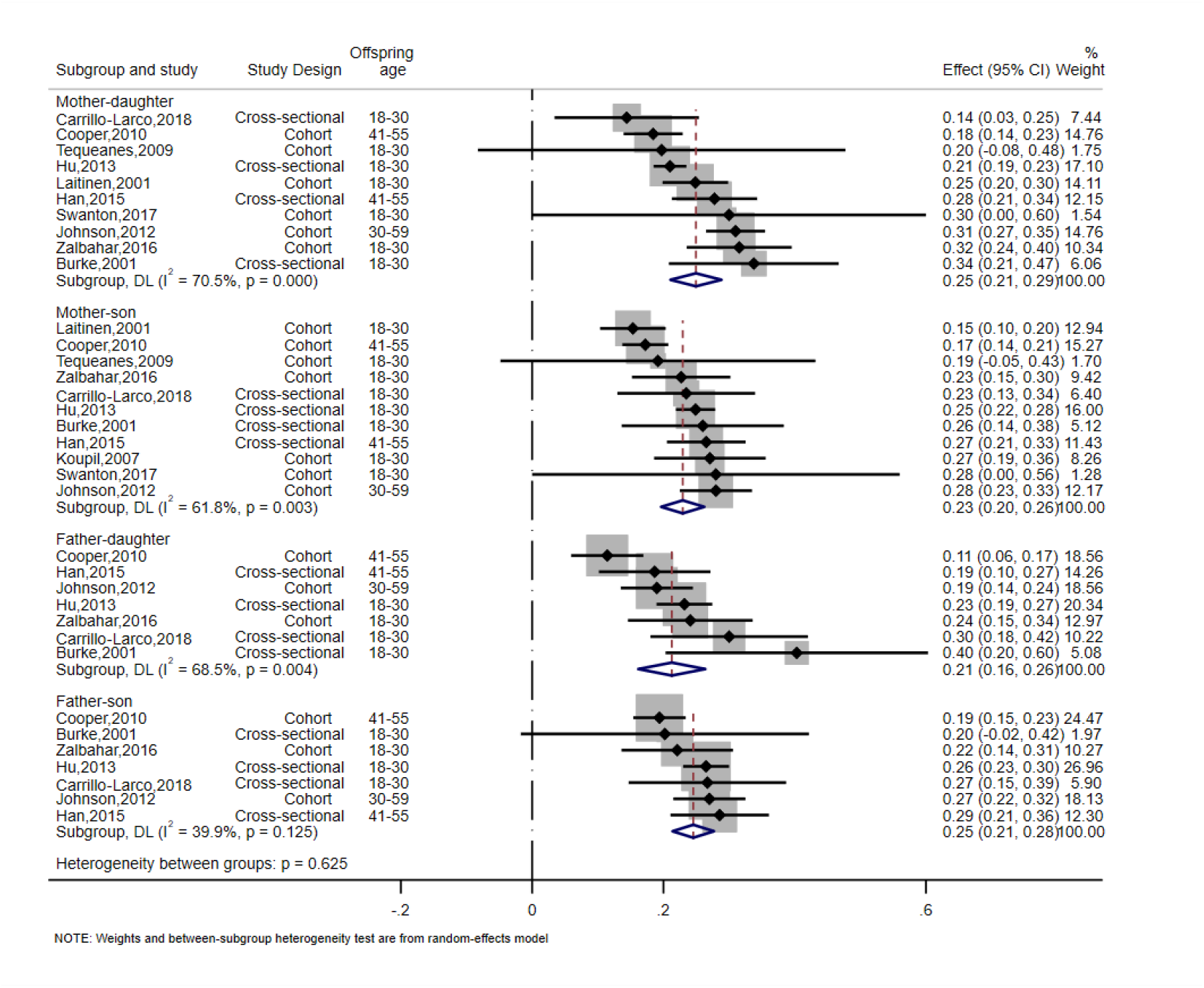
Meta-analysis of the association between parent and offspring BMI at sex-specific level (standardized mean difference) The effect size estimate is the standardized mean difference with 95% confidence intervals (per SD of parental BMI) The size of the squares estimates is proportional to the weight assigned to each study. Diamonds represent pooled estimates from a random effects meta-analysis. The *I*^2^ and *P* values for heterogeneity are shown. BMI, body mass index; CI, confidence interval

Pooled results from 15 studies of the mean difference in offspring BMI (kg/m^2^) per 1 kg/m^2^ parental BMI were consistent with those for the SMD. There were also high levels of between study heterogeneity (*Figure S3a, Figure S3b and Table S6*).

We further compared the strength of associations between maternal and paternal BMI and offspring BMI by calculating the difference in SMD. There was no strong evidence for differences by offspring gender for either maternal or paternal associations (difference in SMD=0.01, 95%CI: -0.02, 0.05; *Figure 3*). Similar approaches were applied to unadjusted SMD and MD, and results remained similar (*Figure S4, Figure S5 and Tables S7 to S9*). There were 4 studies that could not be included in the meta-analyses^33,37,49,50^. The reported findings were consistent with the meta-analysed studies. For detailed descriptions, see *Table S10*.

**Figure 3.**
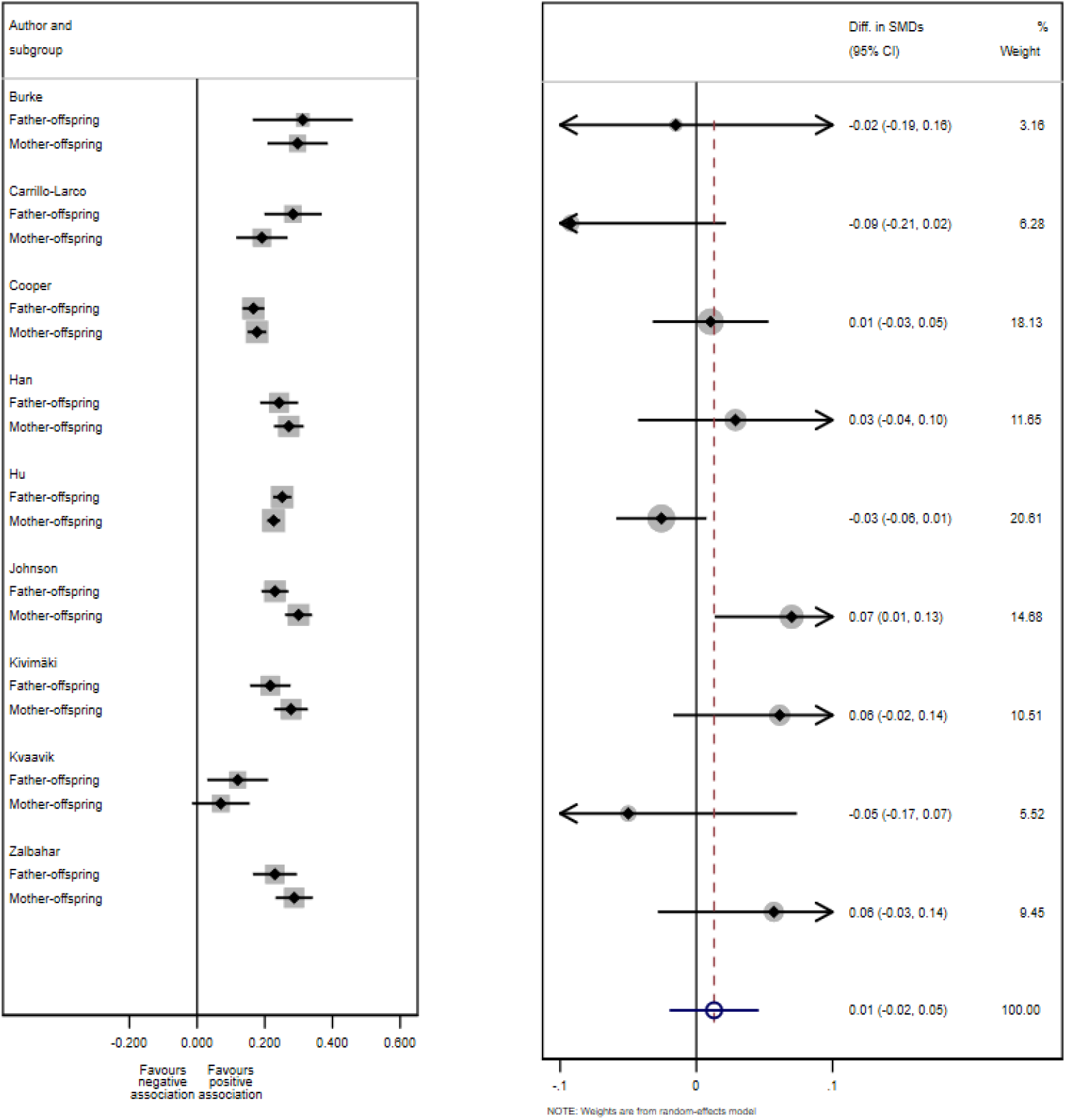
Difference in standardized mean difference between mother-offspring and father-offspring BMI associations Left plot: SMD (95% CI) in each subgroup of each study. Right plot: Difference of SMD comparing effects for mother-offspring and father-offspring in each study with a fixed-effect meta-analysis of these differences of SMD. The size of the squares and circles is proportional to the weight assigned to each study. Open circles represent pooled estimates. SMD, Standardized mean difference; CI, confidence interval

### 3.3 Subgroup and post hoc analyses

There was evidence that associations were weaker when parental BMI was based on parental self-reported weight and height rather than with clinical or research measures (for example SMD=0.21 (95%CI: 0.15, 0.27 for mother-offspring, p=0.000 and SMD=0.26, 95%CI: 0.23,0.30, p=0.006, respectively; (exact p<0.1 for heterogeneity between groups, *Figure S6, Table S11*). The association was higher in cross-sectional studies compared to cohort studies for father-offspring associations but not for mother-offspring associations (*Figure S7a, Table S12*). Pooled results were very similar comparing studies where maternal BMI ascertainment was before/ in early pregnancy or after pregnancy (*Figure S8, Table S13*), similarly results were consistent between studies in which children younger than 30 years and those who were older than 40 years (*Figure S9a, Figure S9b, and Table S14*).

Post hoc analysis were performed to investigate the moderate-to-high degree of between-study heterogeneity, including taking out studies with a small proportion of offspring younger than 18 years (*Table S15*) or with low quality (*Table S16*). The results remained similar to the main analyses.

### 3.4 Secondary outcome - ORs

18 studies investigated the association between parental BMI status and offspring overweight or obesity and were included. 11 of the studies^18,25,31,33,42,47,51-64^ used World Health Organization^65^ categories to define overweight (BMI 25.0–29.9 kg/m^2^) and obese status (BMI ≥30 kg/m^2^); the remaining studies used different thresholds based on the study population distribution.

As noted in the methods we did not pool results from these studies. Most studies found that adult offspring of overweight or obese parents (either mother or father, or both) were more likely to be overweight or obese themselves, compared to those with normal-weight parents, though magnitudes of association varied ranging from 1.1 (95%CI: 0.8, 1.6)^53^ to 11.8 (95%CI: 7.7, 18.0)^18^ (see *Figures S10 to S12*).

Only one study found little evidence of an association between mother-offspring or father-offspring weight status^56^. ORs were generally higher when both parents were overweight or obese compared to results using where only one parent was overweight or obese, and there was evidence of a dose response with stronger magnitudes of association for parental obesity than overweight. 4 studies examined the association of parental BMI as a continuum with ORs for offspring overweight or obesity, with results for theseshowing higher odds or offspring overweight and obesity with higher parental mean BMI^31,62,63,66^.

### 3.5 Study quality and risk of bias

The majority of the studies were considered of good quality (scored above 3), with 4 studies scoring 7, and 3 studies below 2. 11 studies did not have a representative sample of the target population, the exposure or outcome were self-reported or without description in 15 studies, and confounding factors were not considered in the analyses in 10 studies. 13 cross-sectional studies did not score a point for adequacy of follow up of the cohort due to the nature of the study design; 17 cohort studies had a follow-up of <80% and did not provide any description of loss to follow-up. All studies were followed-up long enough for outcomes to occur (average offspring age was above 18 years old). Sensitivity analyses taking out the 3 studies of poor quality (score below 2) showed slightly higher SMD and lower I**^2^** in the adjusted models (*Table S17*).

### 3.6 Funnel plot asymmetry

The funnel plot and Egger’s test shows no strong evidence of funnel asymmetry suggesting there was little evidence of publication bias (*Figure S13*).

## 4. DISCUSSION

In this systematic review, we found positive associations between parental BMI and their offspring BMI in adulthood, assessed using several different measures of association. Directions and magnitudes of associations were similar for both parents and when assessed separately in daughters and sons. They were also similar in subgroups of studies that differed in age at which offspring weight and height were ascertained and timing of maternal BMI in relation to pregnancy. Most studies were considered good quality, with only few scoring poorly, and we observed that results in studies with BMI (parental and/or offspring) based on self-reported rather than measured weight and height were weaker. The findings extend a previous systematic review on the intergenerational transmission of parental and offspring adiposity in childhood^6,19^ and demonstrate that the association persists into adulthood. The magnitude of association across the BMI distribution was relatively weak. For example, the pooled SMD=0.23 per 1SD higher in maternal BMI is equivalent to a correlation coefficient of 0.23. On the other hand, this correlation could be important at a population level.

Most of the meta-analysed studies measured parental BMI concurrently with offspring BMI, and it is conceivable that any intrauterine effects might be diluted with increasing age. However, our results were similar for studies that measured parental BMI before the index pregnancy. That we see similar magnitudes of associations for mothers and fathers, is consistent with previous parental negative control and Mendelian randomization studies^15,16,67-71^, which suggest that intrauterine mechanisms are not key to cross-generational associations. The comparison of same-sex versus cross-sex association also did not reveal significant sex differences in the associations. Our findings are consistent with both parents contributing to an ‘obesogenic’ family environment, but are also compatible with the inheritance from either parent, or both, of genetic traits predisposing to adiposity^72^. Thus, these findings emphasize the need to target all family members for obesity prevention.

Between-study heterogeneity was moderate-to-high for some of the pooled results. We conducted a series of subgroup analyses to explore the sources of heterogeneity, but found consistent results between subgroups for all of these, except whether BMI was based on self-reported or measured weight and height. Results were consistent between studies that did not adjust for any confounders and those adjusting for some confounders (most of which adjusted for what we consider to be key confounders). Thus differences in confounder adjustments are unlikely to explain between study heterogeneity and we are unable to determine the cause of between study heterogeneity. 6 studies adjusted for early life variables such as birth weight or gestational weight gain^40,43,59,61,73,74^, which are potential mediators on the causal pathway between parental and offspring BMI, rather than confounders. Therefore, we used results from models that did not adjust for birthweight or gestational weight gain in the meta-analyses.

Key strengths of our review include the focus on offspring adult BMI, carefully considering which studies are appropriate to meta-analyses, and undertaking relevant subgroup analyses. We have investigated sex-specific associations and compared associations of maternal-offspring to paternal-offspring BMI associations to explore the potential etiology of BMI intergenerational associations.

We acknowledge the limitations of the meta-analyses. Firstly, whilst our focus was on offspring adult (18 years or older) BMI, 7 studies^26,28-33^ (corresponding to 15% of studies and 5% of participants) measuring offspring BMI at 15–25 years were included in the systematic review. However, meta-analysis results were not changed with removal of these studies. We have focused on adult BMI and it is possible that results would be different for other measures of adiposity, such as waist circumference or fat mass. However, the strong correlations between BMI and these measurements, together with similar Mendelian randomization results for maternal BMI with offspring BMI and fat mass, makes this unlikely^75^. The small number of studies reporting all combinations of mother, father, daughter and son associations limited our ability to assess any dose-response relationships between within these different groups.

## Conclusions

This systematic review and meta-analysis of observational studies indicates that the intergenerational associations between parental and offspring obesity persisted into adulthood. We found there was no strong evidence of differences between maternal and paternal lines, which together with Mendelian randomization and negative control studies^6,16^ mostly in younger aged offspring, suggest that intrauterine effects related to higher maternal BMI are not a major cause of higher offspring adiposity.

## Data Availability

All data produced in the present study are available upon reasonable request to the authors

## ACKNOWLEDGEMENT

We thank Mette Lise Lousdal (M.L.L) and Helene Tilma Vistisen (H.T.V) for their support in the literature search and screening.

## Abbreviations

BMI: body mass index
CI: confidence interval
MD: mean difference
OR: odds ratio
RR: risk ratio
ROR: ratio of odds ratio
SMD: standardized mean difference

## AUTHOR CONTRIBUTIONS

JZ and CCD had the original idea for the paper and designed the study. JZ, GC, KO, AO, DAL and CCD contributed to the analysis plan. The bibliographic search and data extraction was carried out by JZ, HTV, MLL, and CCD. The methods and statistical analysis were performed by JZ and GC. The interpretation of results and writing and final editing was done by all authors. All authors reviewed and approved the manuscript.

