## Supplementary Figure1-13 for "Body mass index in parents and their adult offspring: a systematic review and meta-analysis"

**Supplementary Figures**


**Figure S1. PRISMA flow diagram of the study selection process**

**Figure S2a. Meta-analysis of the association between parent and offspring BMI (standardized mean difference)**

**Figure S2b. Meta-analysis of the association between parent and offspring BMI at sex-specific level (standardized mean difference)**

**Figure S3a. Meta-analysis of the association between parent and offspring BMI (mean difference)**

**Figure S3b. Meta-analysis of the association between parent and offspring BMI at sex-specific level (mean difference)**

**Figure S4. Difference of standardized mean difference between mother-offspring and father-offspring association**

**Figure S5. Difference of mean difference between mother-offspring and father-offspring association**

**Figure S6. Standardized mean difference between parent-offspring BMI association-subgroup analyses by BMI measurement methods**

**Figure S7a. Standardized mean difference between parent-offspring BMI association-subgroup analyses by study design**

**Figure S7b. Standardized mean difference between parent-offspring BMI association-subgroup analyses by study design (sex-specific level)**

**Figure S8. Standardized mean difference between parent-offspring BMI association-subgroup analyses by maternal BMI measurement time**

**Figure S9a. Standardized mean difference between parent-offspring BMI association-subgroup analyses by offspring age**

**Figure S9b. Standardized mean difference between parent-offspring BMI association-subgroup analyses by offspring age (sex-specific level)**

**Figure S10. Forest plot showing odds ratio (OR) of offspring being overweight with parental weight status**

**Figure S11. Forest plot showing odds ratio (OR) of offspring being obese with parental weight status**

**Figure S12. Forest plot showing odds ratio (OR) of offspring being overweight or obese with parental weight status**

**Figure S13. Funnel plot for publication bias**

**Figure S1. PRISMA flow diagram of the study selection process**

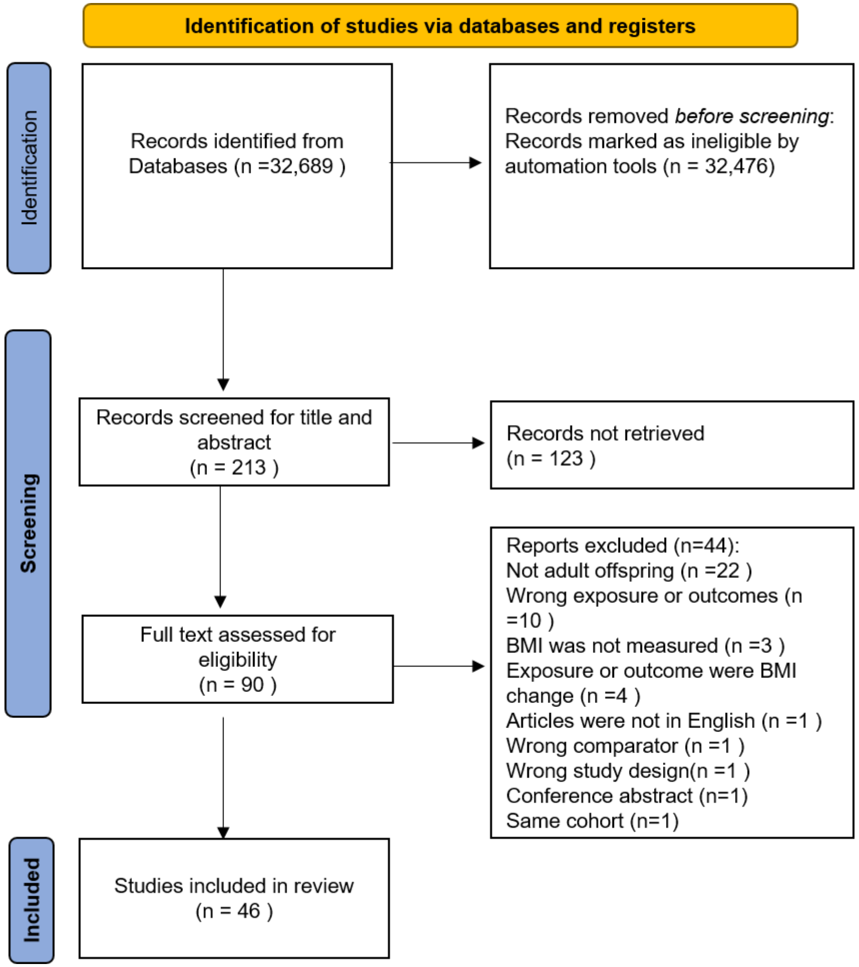

**Figure S2a. Meta-analysis of the association between parent and offspring BMI (standardized mean difference)***

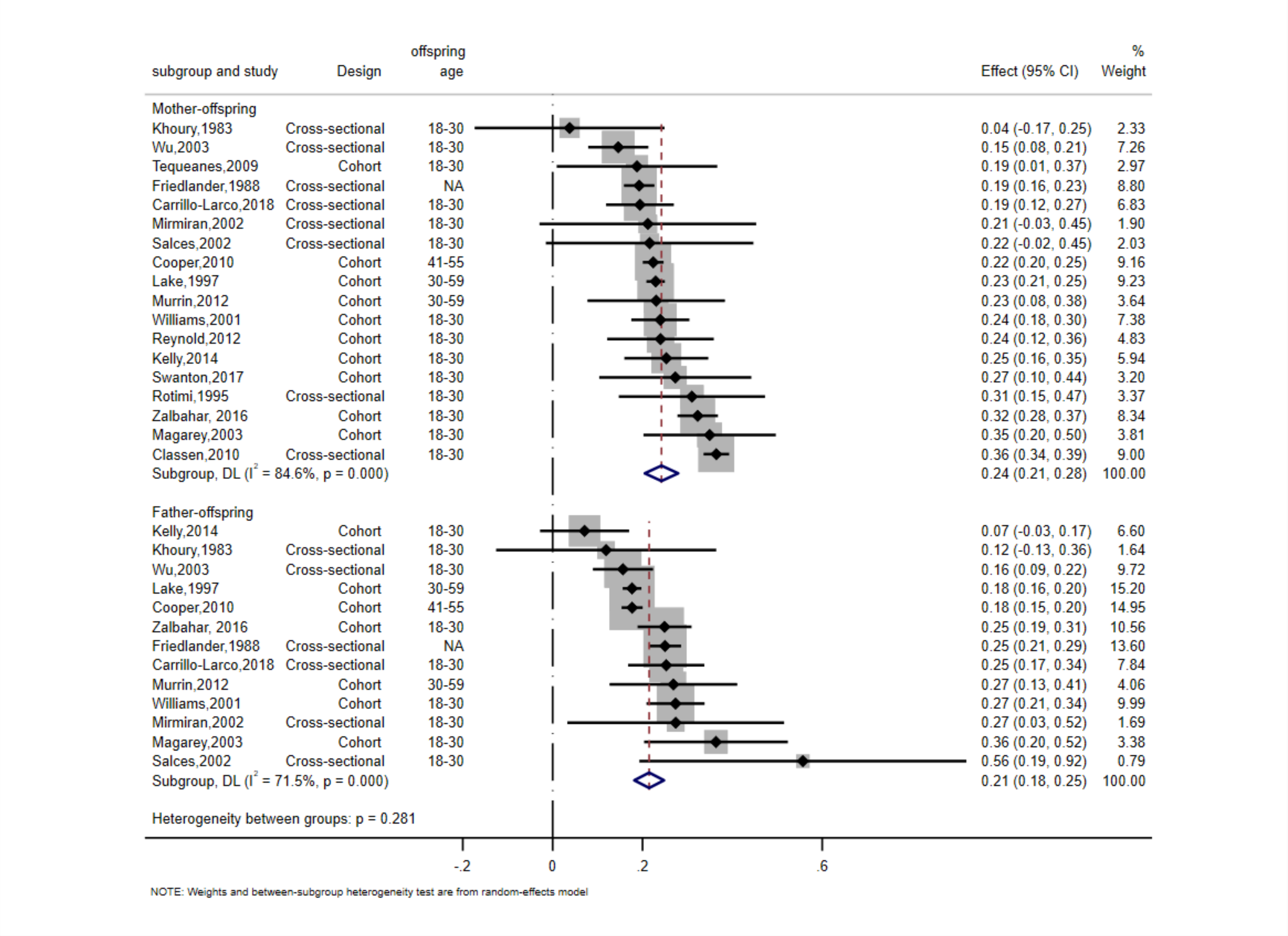


***pooled SMD for unadjusted models.

### Figure S2b. Meta-analysis of the association between parent and offspring BMI at sex-specific level (standardized mean difference)*

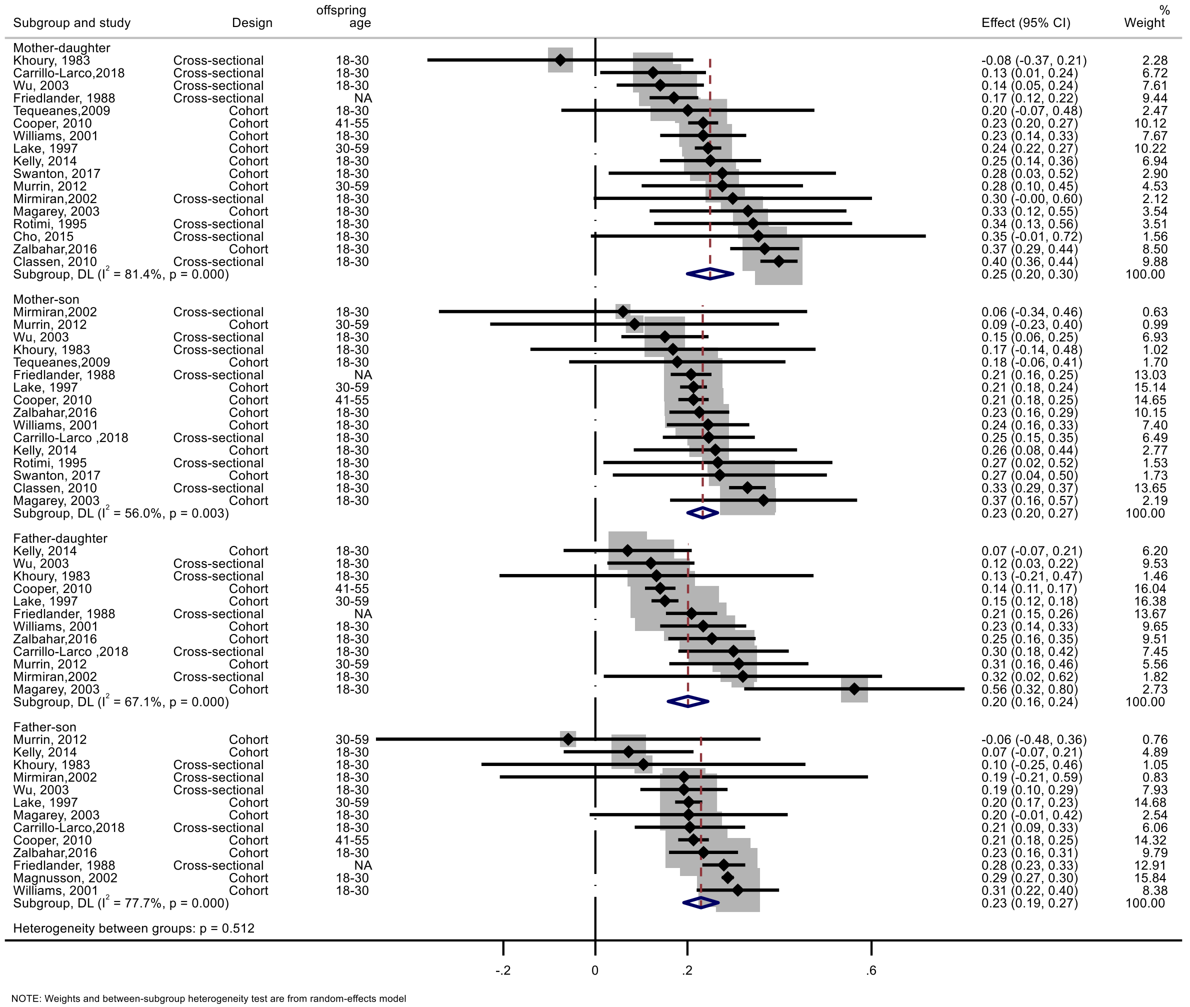


*pooled SMD for unadjusted models

**Figure S3a. Meta-analysis of the association between parent and offspring BMI (mean difference)**

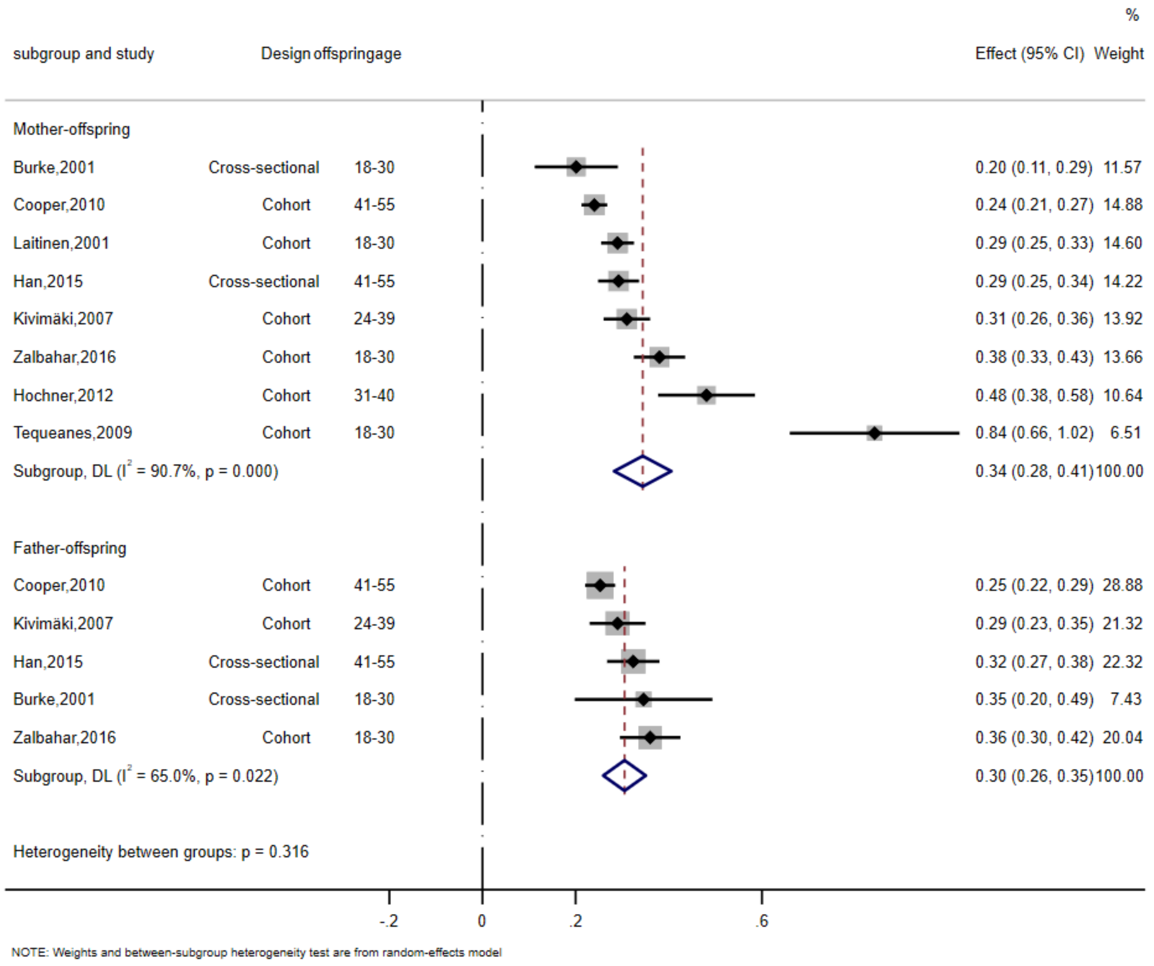

**Figure S3b. Meta-analysis of the association between parent and offspring BMI at sex-specific level (mean difference)**

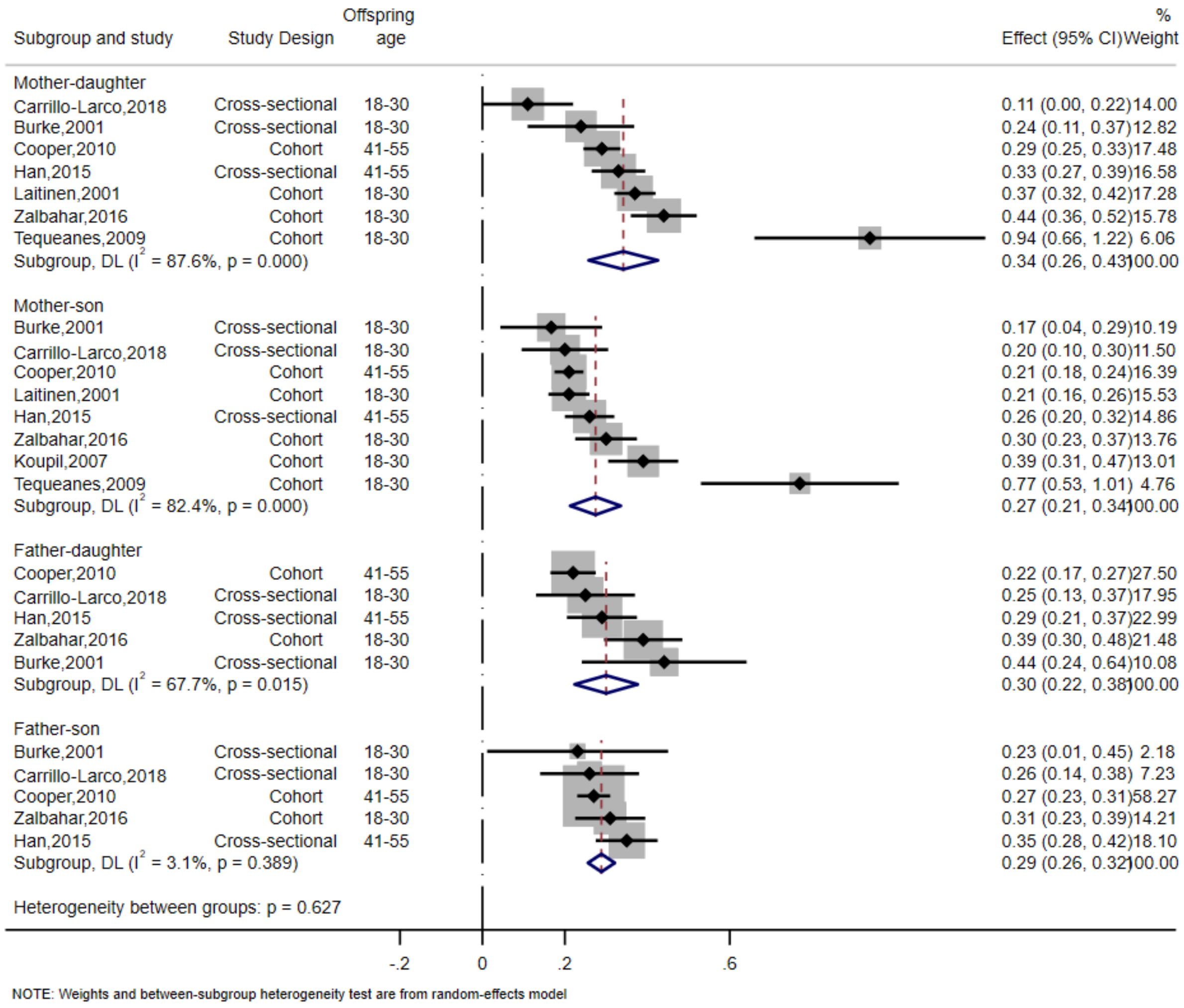

**Figure S4. Difference of standardized mean difference between mother-offspring and father-offspring association***

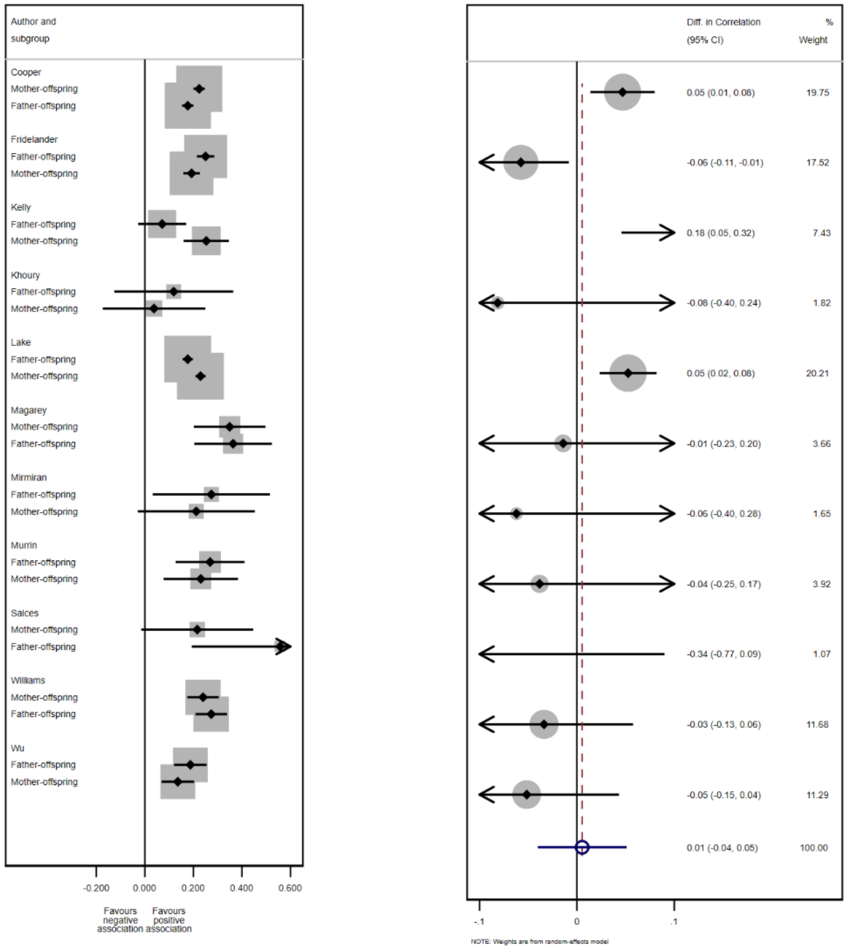


*unadjusted models

**Figure S5. Difference of mean difference between mother-offspring and father-offspring association**

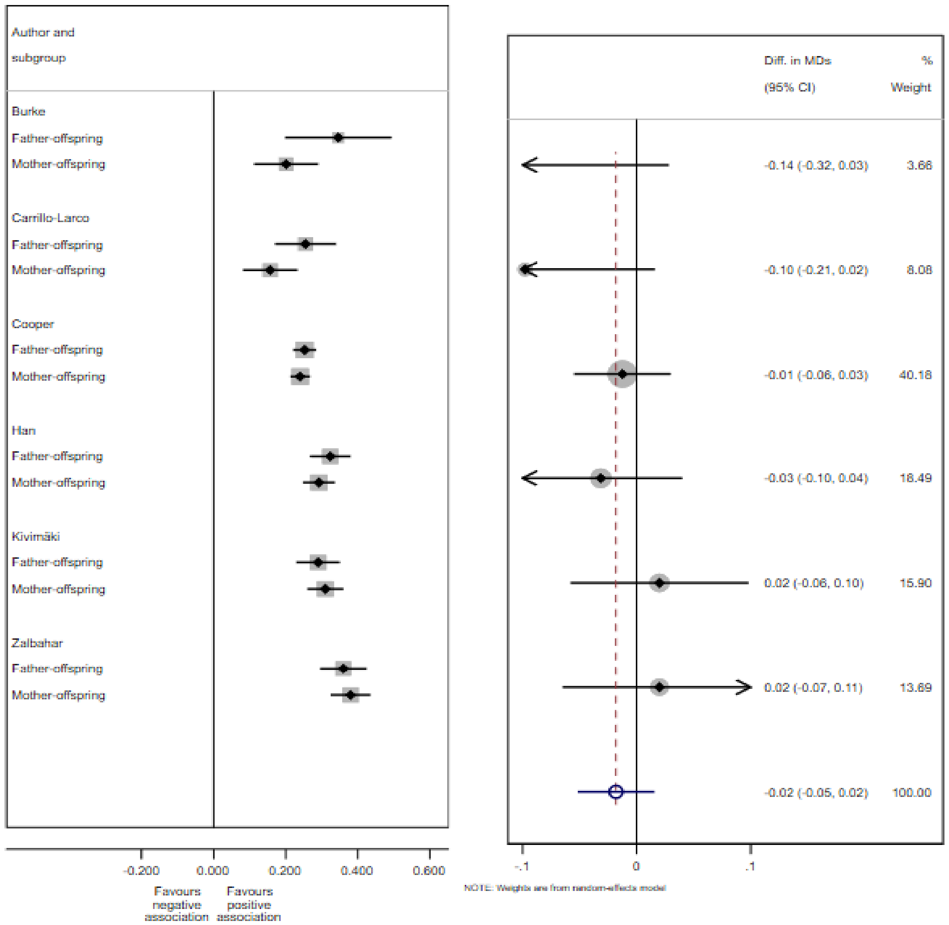

**Figure S6. Standardized mean difference between parent-offspring BMI association-subgroup analyses by BMI measurement methods**

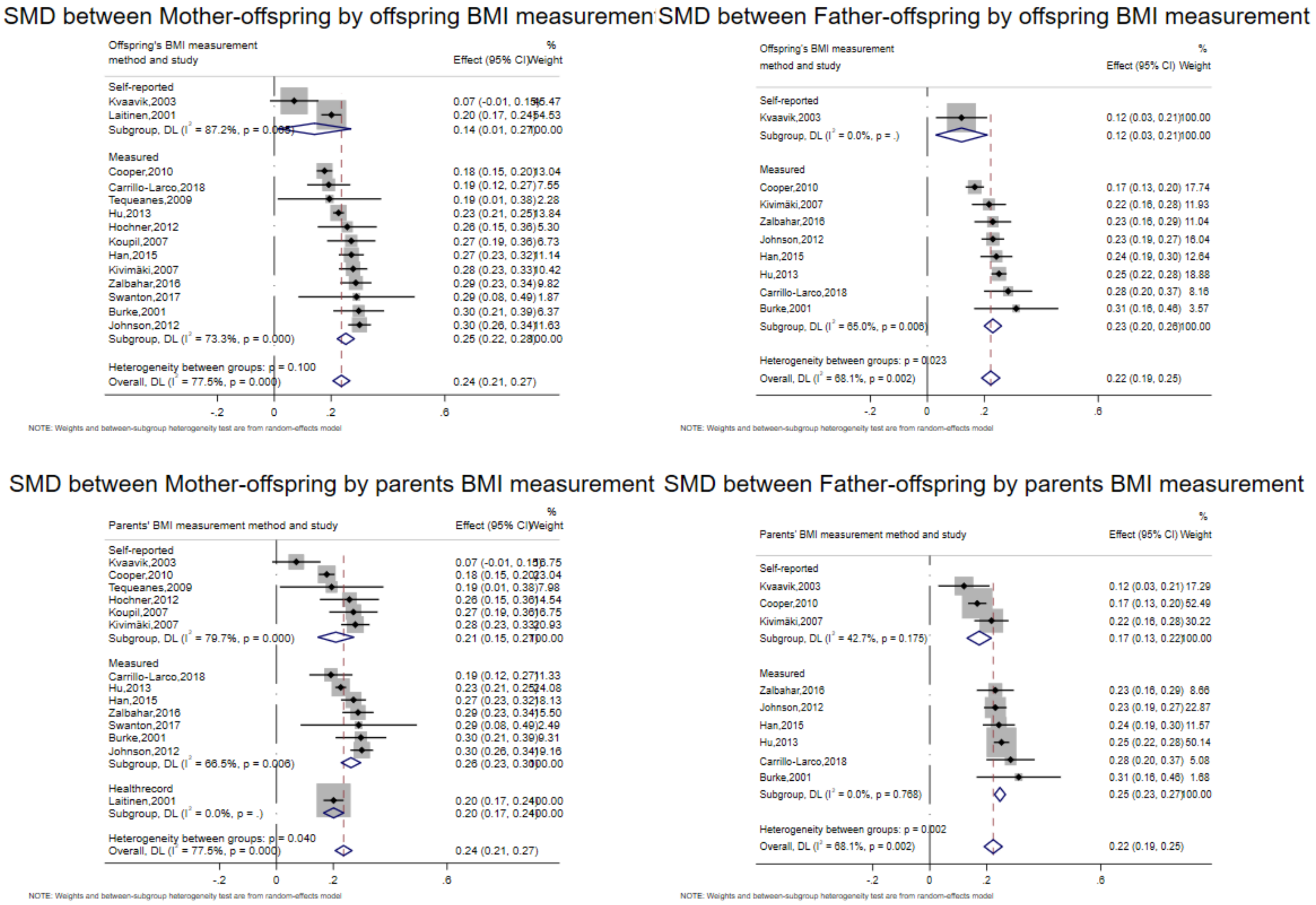


SMD-Standardized mean difference

**Figure S7a. Standardized mean difference between parent-offspring BMI association-subgroup analyses by study design**

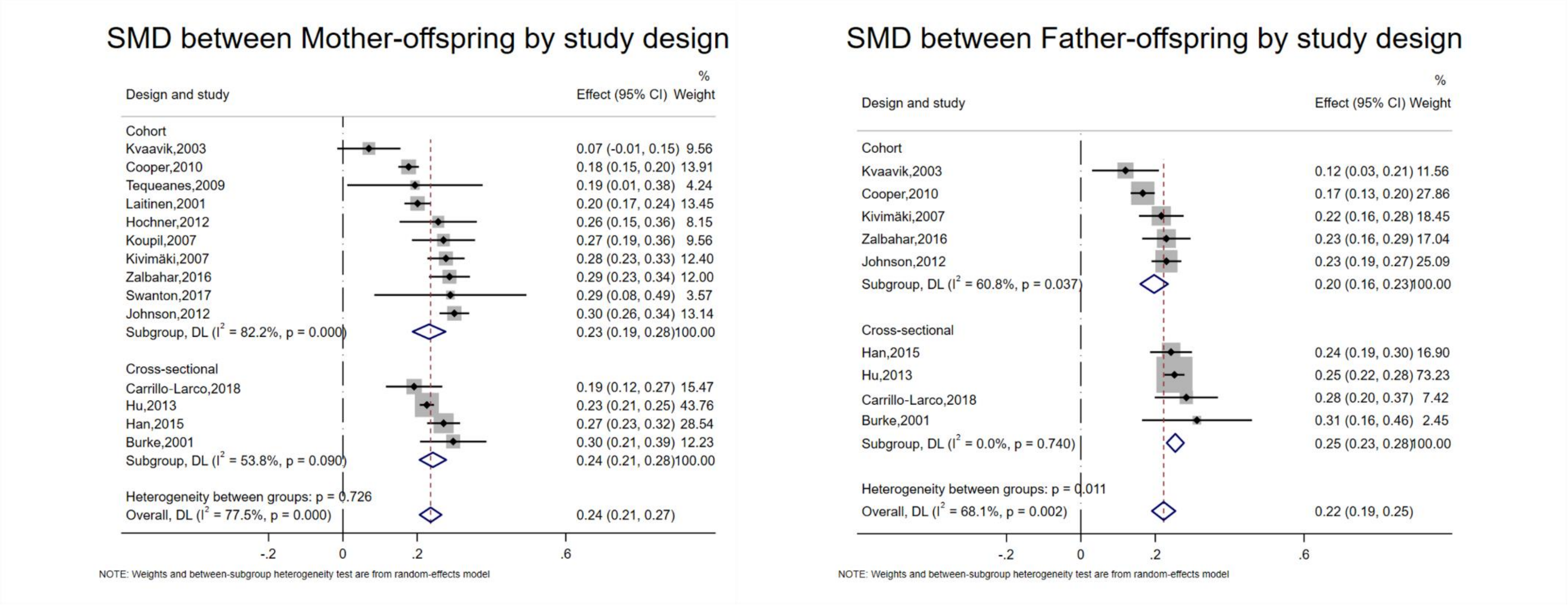


SMD-standardized mean difference

**Figure S7b. Standardized mean difference between parent-offspring BMI association-subgroup analyses by study design (sex-specific level)**

SMD between Mother-daughter by study design

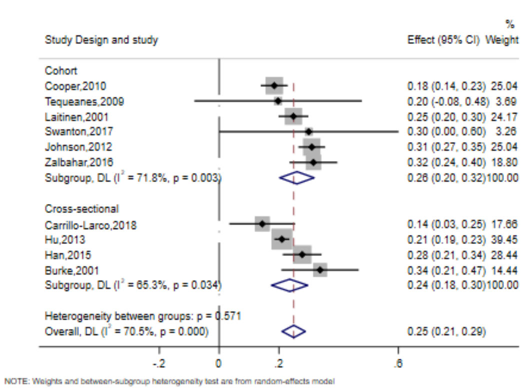


SMD between Mother-son by study design

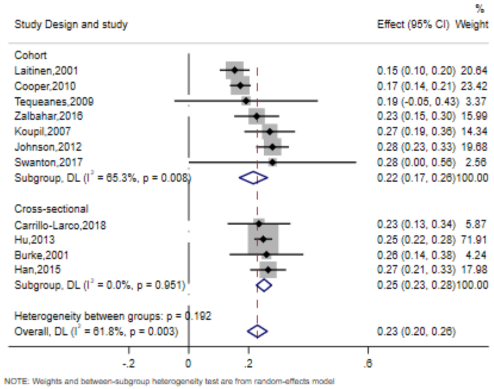


SMD between Father-daughter by study design

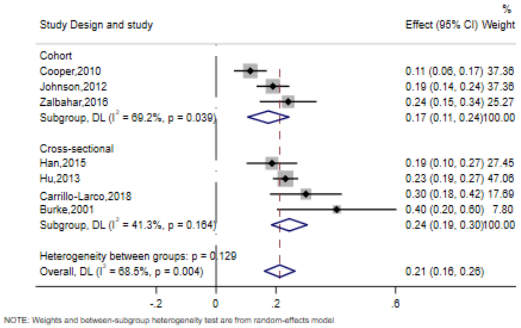


SMD between Father-son by study design

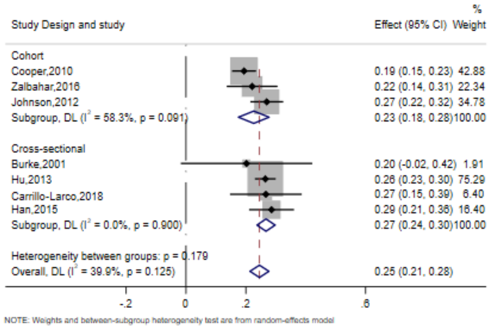


SMD-standardized mean difference

**Figure S8. Standardized mean difference between parent-offspring BMI association-subgroup analyses by maternal BMI measurement time**

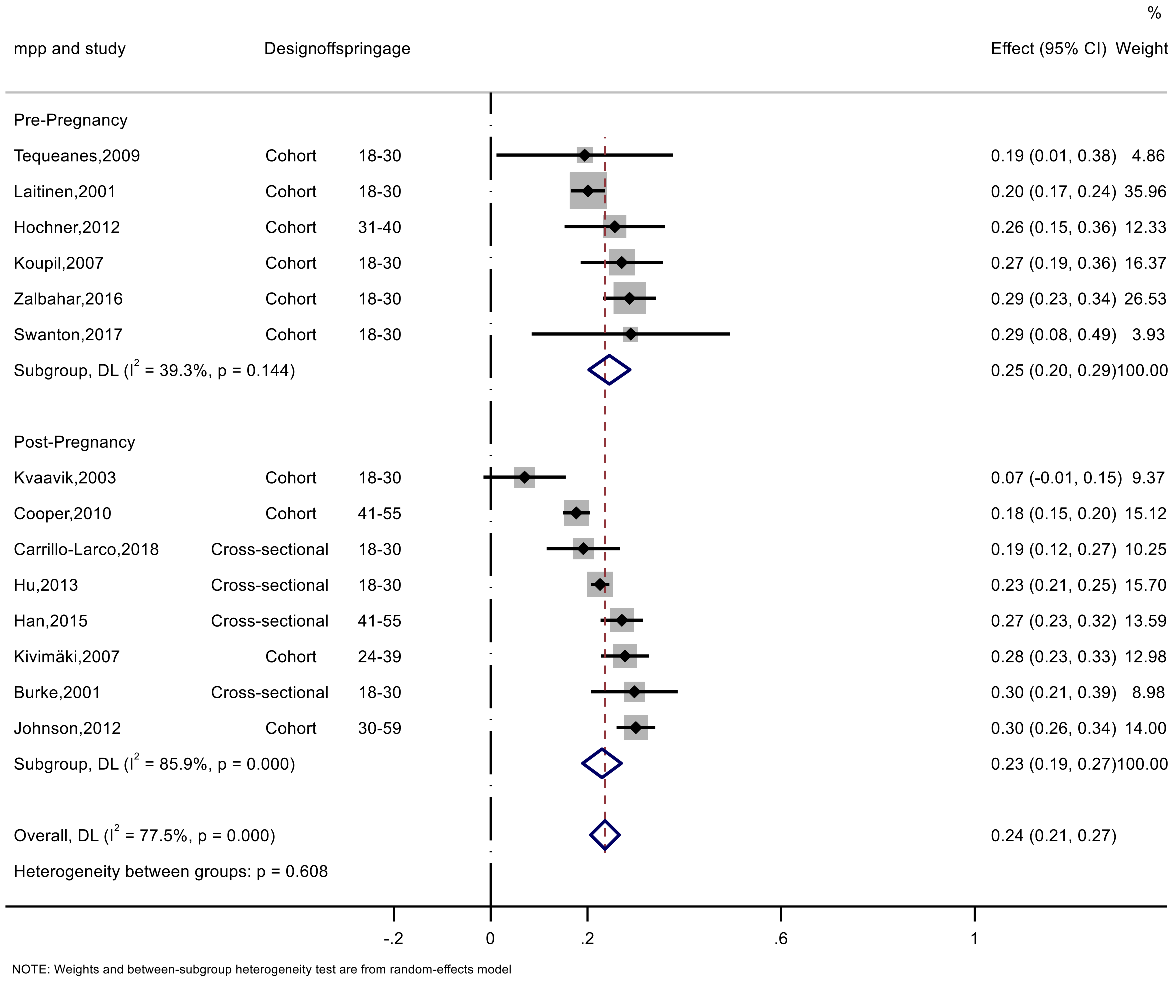


SMD-standardized mean difference; mpp, maternal pre-pregnancy BMI

**Figure S9a. Standardized mean difference between parent-offspring BMI association-subgroup analyses by offspring age**

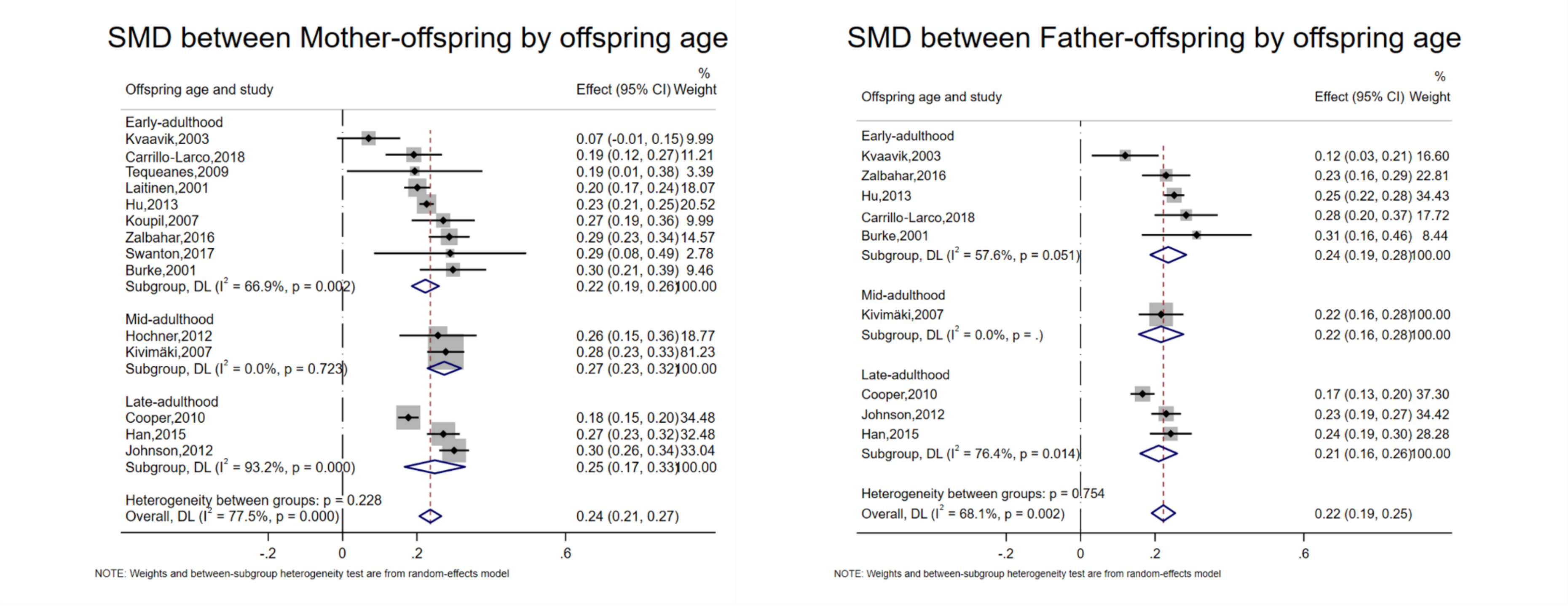


SMD- Standardized mean difference

*Early adulthood: 18-30y; Mid-adulthood: 25-39y or 30-40y; Late adulthood: >40y

**Figure S9b. Standardized mean difference between parent-offspring BMI association-subgroup analyses by offspring age (sex-specific level)**

SMD between Mother-daughter by offspring age

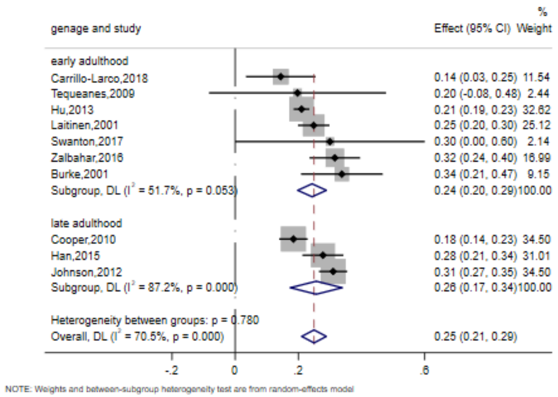


SMD between Mother-son by offspring age

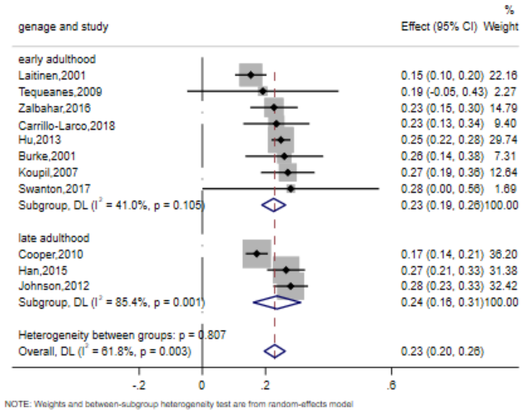


SMD between Father-daughter by offspring age

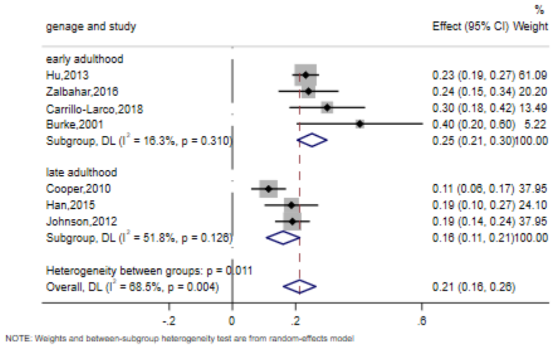


SMD between Father-son by offspring age

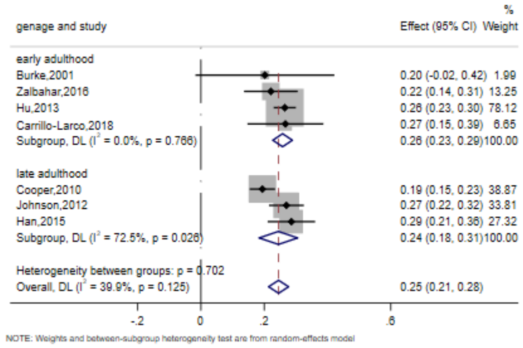


SMD-standardized mean difference

**Figure S10. Forest plot showing odds ratio (OR) of offspring being overweight with parental weight status**

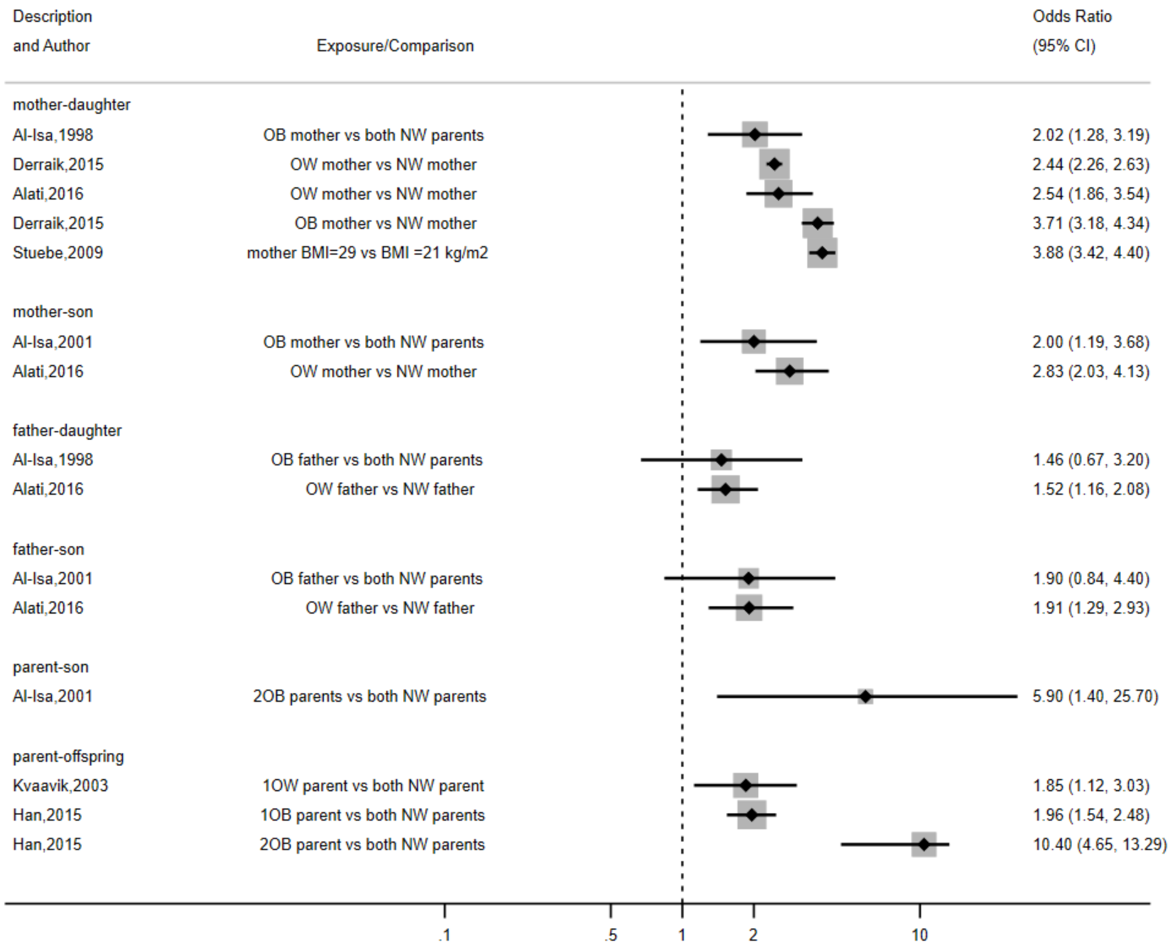


OB-obesity, OW-overweight, NW, normal weight

**Figure S11. Forest plot showing odds ratio (OR) of offspring being obese with parental weight status**

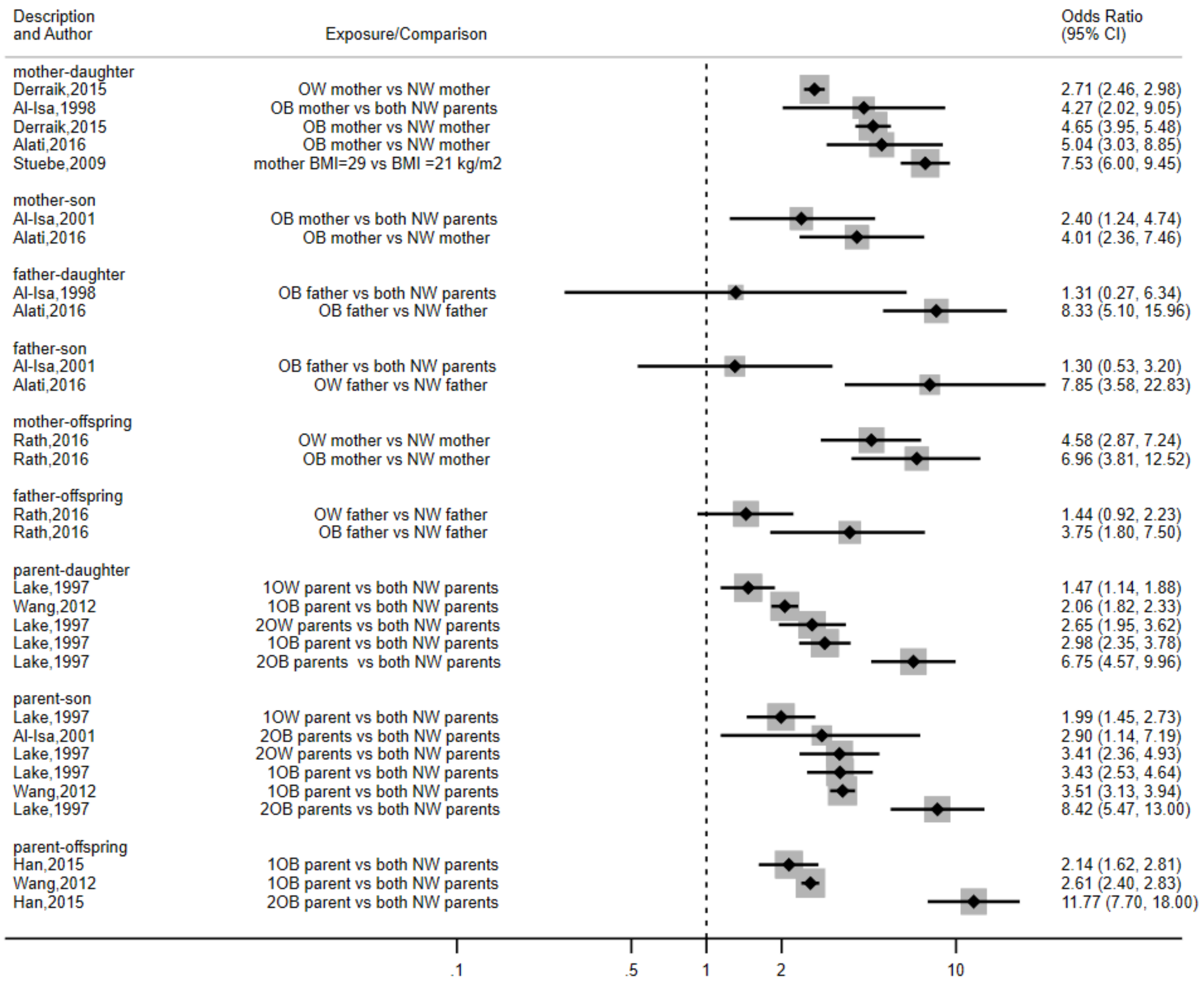


OB-obesity, OW-overweight, NW, normal weight

**Figure S12. Forest plot showing odds ratio (OR) of offspring being overweight or obese with parental weight status**

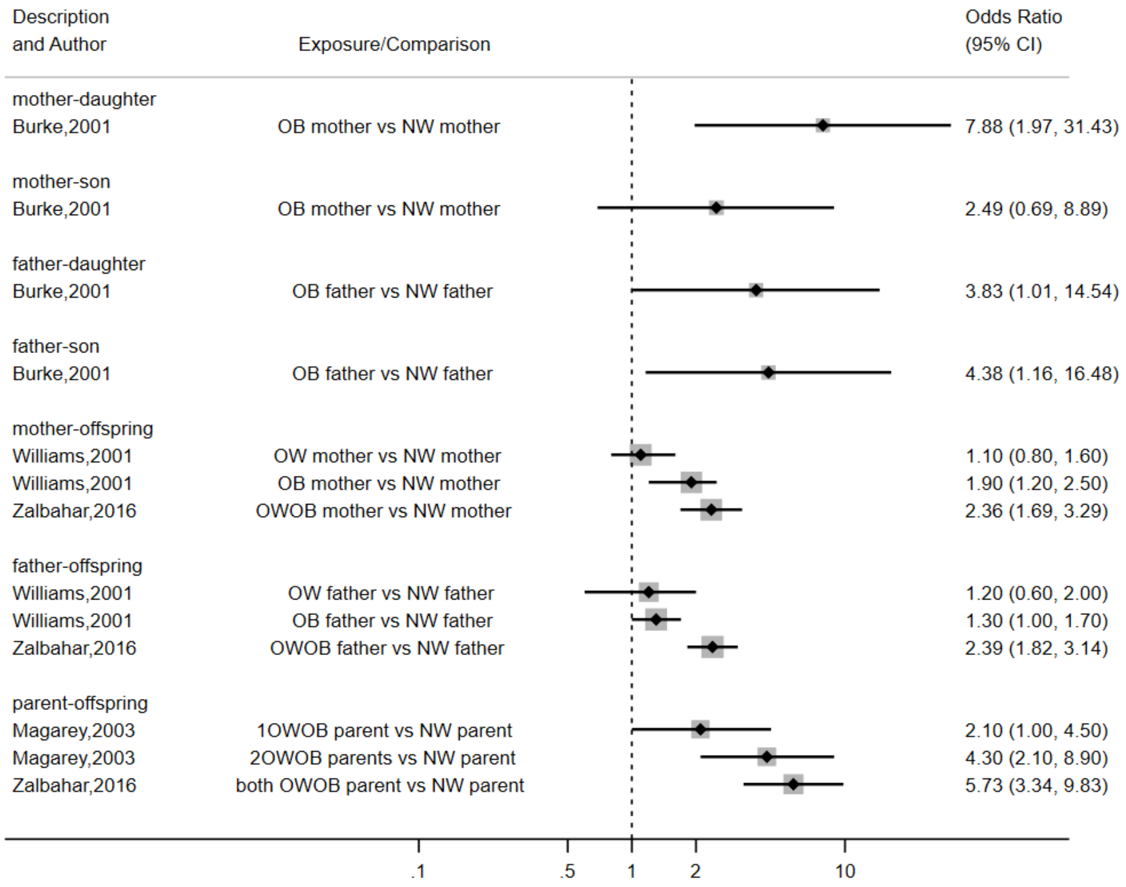


OB-obesity, OW-overweight, NW, normal weight, OWOB, overweight or obese

**Figure S13. Funnel plot for publication bias**

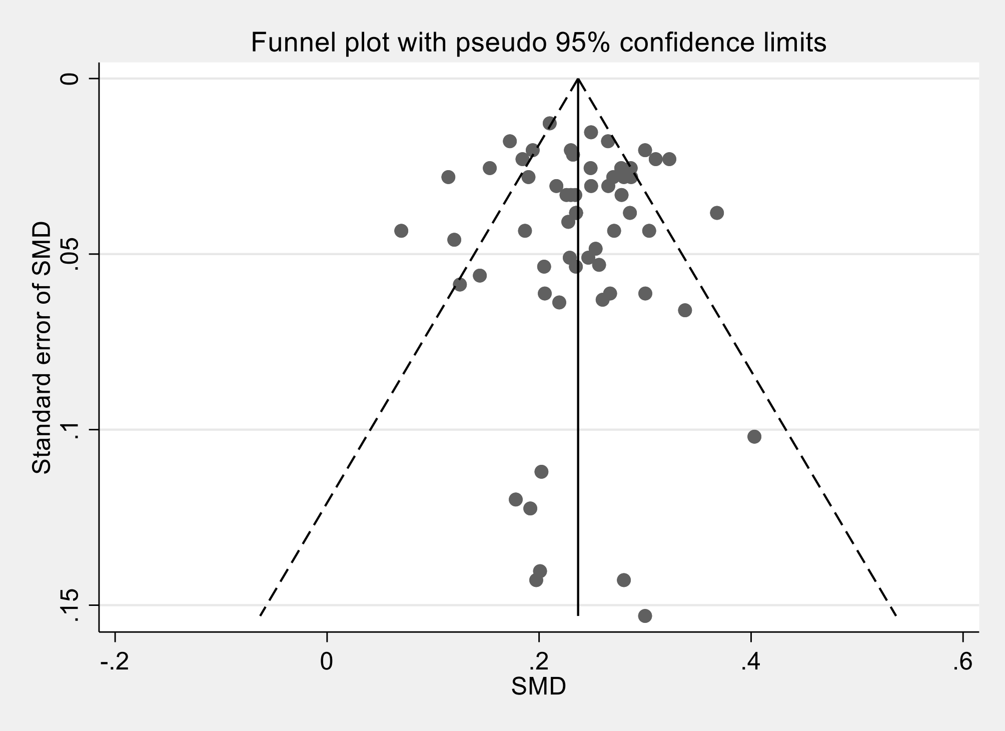
