## Supplementary Table 1-17 for "Body mass index in parents and their adult offspring: a systematic review and meta-analysis"

### Contents


**Table S1. Description of all the studies included in systematic review and meta-analyses**

**Table S2. Descriptions of studies reporting correlation coefficient**

**Table S3. Descriptions of studies reporting mean difference or standardized mean difference**

**Table S4. Descriptions of studies reporting odd ratios or risk ratios**

**Table S5. Pooled standardized mean difference between parental and offspring BMI (per SD)**

**Table S6. Pooled mean difference between parental and offspring BMI (per kg/$m^2$)**

**Table S7. Difference of standardized mean difference between maternal and paternal line in adjusted models**

**Table S8. Difference of standardized mean difference between maternal and paternal line in unadjusted models**

**Table S9. Difference of mean difference between maternal and paternal line**

**Table S10. Summary of studies not included in meta-analyses**

**Table S11. Subgroup analyses by BMI measurement method**

**Table S12. Subgroup analyses by study design**

**Table S13. Subgroup analyses by maternal BMI measurement time**

**Table S14. Subgroup analyses by offspring age**

**Table S15. Post hoc analyses-standardized mean difference taking out studies including younger participants**

**Table S16. Post hoc analyses-standardized mean difference taking out studies with low quality score**

**Table S17. Study quality assessment using Adapted Newcastle-Ottawa scale**

**Table S1. Description of all the studies included in systematic review and meta-analyses**

| Author, publication year, country | Title | Data source | Study design | Sample size | Parents age (years, mean(sd)) | Offspring age (years,mean(sd)) | Assessment of anthropometrics | Parental variable(s) | Offspring variable(s) | Reported measure-ment of association |
|---|---|---|---|---|---|---|---|---|---|---|
| Khoury et al.[1] 1983 USA | Parent-offspring and sibling body mass index associations during and after sharing of common household environments: the Princeton School District Family Study | The Princeton School Family Study cohort | Cross-sectional study | 877 | NA | Above 20 | NA | the Quetelet index (weight/ height2), and the Benn index (weight/height$^2$) | the Quetelet index (weight/ height$^2$), and the Benn index (weight/height$^2$) | correlation coefficient |
| Friedlander et al.[2] 1988 Israel | Familial aggregation of body mass index in ethnically diverse families in Jerusalem | The Jerusalem LRC study population | Cross-sectional study | 2,942 nuclear family | NA | Above 17 | M:Measured<br>F: Measured<br>C: Measured | BMI continuous | BMI continuous | correlation coefficient |
| Sørensen et al.[3] 1992 Denmark | Correlations of body mass index of adult adoptees and their biological and adoptive relatives | The non-familial adoptions granted in the Copenhagen area | Cross-sectional study | 3,476 | NA | 42.2 (8.1) | NA | BMI continuous | BMI continuous | correlation coefficient |

| Author, publication year, country | Title | Data source | Study design | Sample size | Parents age (years, mean(sd)) | Offspring age (years,mean(sd)) | Assessment of anthropometrics | Parental variable(s) | Offspring variable(s) | Reported measure-ment of association |
|---|---|---|---|---|---|---|---|---|---|---|
| Rotimi et al.[4] 1995 USA | Familial resemblance for anthropometric measurements and relative fat distribution among African Americans. International journal of obesity and related metabolic disorder | Survey sample from Maywood, IL | Cross-sectional study | 162 | M:46.8(12.50) F:49.10(12.40) | D: 25.40(7.80) S: 26.60(6.30) | M:Measured F: Measured C: Measured | BMI, WHcR, WC, HC as continuous | BMI, WC, HC, WHR as continuous | correlation coefficient |
| Lake et al.[5] 1997 UK | Child to adult body mass index in the 1958 British birth cohort: associations with parental obesity | The 1958 British birth cohort | Cohort study | 12,747 | When children were aged 11 years | 23 AND 33 [Ø] | M:Self-reported F:Self-reported (at offspring age 11) C:Measured (33 y) | Underweight, Normal, Overweight, or obese, as defined by the 85th centile &continuous | Binary: Obese vs normal &continuous | ORs correlation coefficient |
| Al-Isa et al.[6] 1998 Kuwaiti | Factors associated with overweight and obesity among Kuwaiti college women | Random sample from Kuwaiti college women | Cross-sectional study | 585 | NA | 18-23 | M: Measured F: Self-reported C: Measured | WHO criteria: Underweight Normal Overweight Obese | Binary: Overweight or Obese vs normal | ORs |

| Author, publication year, country | Title | Data source | Study design | Sample size | Parents age (years, mean(sd)) | Offspring age (years,mean(sd)) | Assessment of anthropometrics | Parental variable(s) | Offspring variable(s) | Reported measure-ment of association |
|---|---|---|---|---|---|---|---|---|---|---|
| Burke et al.[7] 2001 Australia | Family lifestyle and parental body mass index as predictors of body mass index in Australian children: a longitudinal study | A cohort of Perth children | Cross-sectional study | 219 families (104 sons, and 115 daughters) | F:52.1 (0.4) M:49.1(0.3) | S: 18.1(0.01) D: 18.1(0.01) | M: Measured F: Measured C: Measured | The National Heart Lung and Blood Institute: Normal Overweight Obesity | OWOB (BMI>25 $kg/m^2$) vs BMI <25 $kg/m^2$ | β (per $kg/m^2$) ORs |
| Williams et al.[8] 2001 New Zealand | Overweight at age 21: the association with body mass index in childhood and adolescence and parents' body mass index. A cohort study of New Zealanders born in 1972–1973 | Birth cohort born in Dunedin, New Zealand | Cohort study | 924 | when offspring was aged 11 years | 21 | M: Self-reported F: Self-reported C: Measured | WHO criteria: Underweight Normal Overweight Obese | OWOB defined as BMI>25 $kg/m^2$ Normal defined as BMI <25 $kg/m^2$ & continuous | RRs correlation coefficient |
| Laitinen et al.[9] 2001 Finland | Family social class, maternal body mass index, childhood | The northern Finland Birth Cohort for 1966 | Cohort study | 6,280 | M: around 27 at antenatal visit | S:25.2 (3.6) D: 23.8(4.4) | M: health record C: Self-reported | BMI continuous | WHO criteria: Underweight Normal Overweight Obese &continuous | β (per $kg/m^2$) |

| Author, publication year, country | Title | Data source | Study design | Sample size | Parents age (years, mean(sd)) | Offspring age (years,mean(sd)) | Assessment of anthropometrics | Parental variable(s) | Offspring variable(s) | Reported measure-ment of association |
|---|---|---|---|---|---|---|---|---|---|---|
| | body mass index, and age at menarche as predictors of adult obesity | | | | | | | | | |
| Magnusson et al.[10] 2002 Sweden | Familial resemblance of body mass index and familial risk of high and low body mass index. A study of young men in Sweden | The Swedish Multi-Generation Register | Cohort study | 22,517 | 18-19 | 18-19 | F: measured | BMI continuous | BMI continuous | correlation coefficient |
| Salces et al.[11] 2002 Spain | Family resemblance for anthropometric traits II. Assessment of maternal occupational and age effects | Nuclear families from the province of Biscay | Cross-sectional study | 1,326 | F:22-66<br>M:22-62 | after puberty | M: Measured<br>F: Measured<br>C: Measured | BMI continuous | BMI continuous | correlation coefficient |
| Mirmiran et al.[12] 2002 Iran | Familial clustering of obesity and the role of nutrition: Tehran Lipid and Glucose | The Tehran Lipid and Glucose Study (TLGS) | Cross-sectional study | 474 | M:40.6 (0.5)<br>F:47.4 (0.5) | 18-25 | M: Measured<br>F: Measured<br>C: Measured | Overweight $\geqslant$25 kg/m$^2$ | Overweight $\geqslant$25 kg/m$^2$ (aged$\geqslant$20) | correlation coefficient ORs*** |

| Author, publication year, country | Title | Data source | Study design | Sample size | Parents age (years, mean(sd)) | Offspring age (years,mean(sd)) | Assessment of anthropometrics | Parental variable(s) | Offspring variable(s) | Reported measure-ment of association |
|---|---|---|---|---|---|---|---|---|---|---|
| | Study. International journal of obesity | | | | | | | | | |
| Kvaavik et al.[13] 2003 Norway | Predictors and tracking of body mass index from adolescence into adulthood: follow-up of 18 to 20 years in the Oslo Youth Study | The Oslo Youth Study | Cohort study | 485 | F: 45.8 (7.0)<br>M: 42.3 (6.3)<br>[when offspring were aged 13 years] | 32.4(1.0) | M: Self-reported<br>F: Self-reported<br>C: Self-reported | BMI⩾25 $kg/m^2$ vs BMI<25 $kg/m^2$ | BMI⩾25 $kg/m^2$ Vs BMI<25 $kg/m^2$ &continuous | β (per sd) ORs |
| Wu et al.[14] 2003 Taiwan | Familial resemblance of adiposity-related parameters: results from a health check-up population in Taiwan | The Mei-Jou Health Screening program | Cross-sectional study | 1,724 | F: 58.2 (8.5)<br>M: 55.3 (8.5) | S: 30.1 (8.2)<br>D: 28.5 (7.2) | M:Measured<br>F: Measured<br>C: Measured | BMI as continuous | BMI as continuous | correlation coefficient |
| Magarey et al.[15] 2003 Australia | Predicting obesity in early adulthood from childhood and parental obesity | Sample selected from healthy term infants born in Adelaide, | Cohort study | 188 | When offspring were aged 8 years | 20 | M: Measured<br>F: Measured<br>C: Measured | Acceptable weight <25 $kg/m^2$, overweight ⩾ 25 and <30 $kg/m^2$ and obese ⩾ 30 $kg/m^2$ | Acceptable weight <25 $kg/m^2$, overweight ⩾ 25 and <30 $kg/m^2$ and obese ⩾ 30 $kg/m^2$ | correlation coefficient ORs |

| Author, publication year, country | Title | Data source | Study design | Sample size | Parents age (years, mean(sd)) | Offspring age (years,mean(sd)) | Assessment of anthropometrics | Parental variable(s) | Offspring variable(s) | Reported measure-ment of association |
|---|---|---|---|---|---|---|---|---|---|---|
| | | South Australia | | | | | | | | |
| Kazumi et al.[16] 2005 Japan | Associations of middle-aged mother's but not father's body mass index with 18-year-old son's waist circumferences, birth weight, and serum hepatic enzyme levels | Male college students from Kobe University of Mercantile Marine | Cross-sectional study | 195 | F: 49 (4)<br>M: 46 (3) | 18 | M: Self-reported<br>F: Self-reported<br>C: Measured | BMI continuous | BMI continuous | Correlation coefficient |
| Crossman et al.[17] 2006 USA | The family environment and American adolescents' risk of obesity as young adults. | The United States National Longitudinal Study of Adolescent Health | Cross-sectional study | 6,378 | NA | 18-26 y | M: Self-reported<br>F: Self-reported<br>C: Self-reported | WHO criteria:<br>Underweight<br>Normal<br>Overweight<br>Obese | CDC's adult guidelines<br>Binary: OW/OB | ORs |
| Kivimäki et al.[18] 2007 Finland | Substantial intergenerational increases in body mass index are not explained by the fetal | The Young Finns Study | Cohort study | 1,918 | F: 40.0 (8.4)<br>M: 37.5 (7.6)<br>[when offspring were aged 3-18y] | 24-39 y | M: Self-reported<br>F: Self-reported<br>C: Measured | BMI continuous | BMI continuous | β (per kg/m$^2$) |

| Author, publication year, country | Title | Data source | Study design | Sample size | Parents age (years, mean(sd)) | Offspring age (years,mean(sd)) | Assessment of anthropometrics | Parental variable(s) | Offspring variable(s) | Reported measure-ment of association |
|---|---|---|---|---|---|---|---|---|---|---|
| | overnutrition hypothesis: the Cardiovascular Risk in Young Finns Study | | | | | | | | | |
| Abu-Rmeileh NM et al.[19] 2008 Scotland | Contribution of Midparental BMI and other determinants of obesity in adult offspring | The Renfrew and Paisley | Cohort study | 2,162 | 45-64 | 30-59 | M:Measured<br>F:Measured<br>C:Measured | WHO criteria:<br>Underweight<br>Normal<br>Overweight<br>Obese<br>&the mean of parental BMI | WHO criteria:<br>Underweight<br>Normal<br>Overweight<br>Obese<br>&continuous | β (per kg/$m^2$) |
| Koupil et al.[20] 2008 Sweden | Social and early-life determinants of overweight and obesity in 18-year-old Swedish men | The Uppsala Birth Cohort | Cohort study | 1,103 | NA | 18 (range:18-23) | M: Self-reported<br>S: Measured | WHO criteria:<br>Underweight<br>Normal<br>Overweight<br>Obese<br>&continuous | WHO criteria:<br>Underweight<br>Normal<br>Overweight<br>Obese<br>&continuous | β (per kg/$m^2$)<br>ORs |
| Tequeanes et al.[21] 2009 Brazil | Maternal anthropometry is associated with the body mass index and waist: height ratio of offspring | 1982 Pelotas Birth Cohort Study | Cohort study | 2,978 | NA | 23 | M: Self-reported<br>C: Measured | BMI<br>Continuous | BMI<br>Continuous | β (per kg/$m^2$) |

| Author, publication year, country | Title | Data source | Study design | Sample size | Parents age (years, mean(sd)) | Offspring age (years,mean(sd)) | Assessment of anthropometrics | Parental variable(s) | Offspring variable(s) | Reported measure-ment of association |
|---|---|---|---|---|---|---|---|---|---|---|
| | at 23 years of age | | | | | | | | | |
| Kowalski-Jones et al.[22] 2009 USA | Are you what your mother weighs? Evaluating the impact of maternal weight trajectories on youth overweight | The National Longitudinal Survey of Youth 1979 Cohort | Cross-sectional study | 1,759 | 14-22 and 39-47 | 16-21 | M: Self-reported and measured<br>C: Self-reported and measured | BMI continuous | Overweight: The CDC BMI-for-age | ORs |
| Stuebe et al.[23] 2009 USA | Maternal-recalled gestational weight gain, pre-pregnancy body mass index, and obesity in the daughter | The Nurses' Healthy Study and the Nurses' Mothers' Cohort | Longitudinal study | 26,506 mother-nurse daughter dyads | NA | 25-44 or 36-56 | M: Self-reported<br>D: Self-reported | Pre-pregnancy BMI:<br>21 kg/m$^2$,<br>23 kg/m$^2$,<br>25 kg/m$^2$,<br>27 kg/m$^2$,<br>29 kg/m$^2$ | Overweight as BMI⩾25 and <30 kg/m$^2$<br>obese as BMI⩾30 kg/m$^2$ | ORs |
| Classen et al.[24] 2010 USA | Measures of the intergenerational transmission of body mass index between mothers and their children in the United | The National Longitudinal Survey of Youth 1979 (NLSY79) and the Children and Young Adults of | Cross-sectional study | 4,748 | 16-24 | 16-24 | M: Measured<br>C: Measured | WHO criteria:<br>Underweight<br>Normal<br>Overweight<br>Obese | BMI continuous | correlation coefficient |

| Author, publication year, country | Title | Data source | Study design | Sample size | Parents age (years, mean(sd)) | Offspring age (years,mean(sd)) | Assessment of anthropometrics | Parental variable(s) | Offspring variable(s) | Reported measure-ment of association |
|---|---|---|---|---|---|---|---|---|---|---|
| | States, 1981–2004 | the NLSY79 (YA NLSY79) | | | | | | | | |
| Cooper et al.[25] 2010 UK | Associations between parental and offspring adiposity up to midlife: the contribution of adult lifestyle factors in the 1958 British Birth Cohort Study | The 1958 British birth cohort | Cohort study | 9,346 | when offspring were aged 11 years | 44-45 | M:Self-reported F:Self-reported (when offspring were aged 11) C:Measured | WHO criteria: Underweight Normal Overweight Obese | BMI continuous | β (per $kg/m^2$) correlation coefficient ORs |
| Reynolds et al.[26] 2010 UK | Maternal BMI, parity, and pregnancy weight gain: influences on offspring adiposity in young adulthood | The Motherwell birth chort study | Cohort study | 276 | 27.2 (6.0) | 27-30 | M: antenatal records C: Measured | BMI continuous | BMI continuous AND binary outcome: overweight (BMI >25$kg/m^2$ ) | correlation coefficient ORs |
| Al-Isa et al.[27] 2011 Kuwaiti | Factors associated with overweight and obesity among Kuwaiti men | Kuwaiti men, samples from ambulatory patients visiting a | Cross-sectional study | 464 | NA | Above 20 | M: Reported F: Reported C: Measured | WHO criteria: Underweight Normal Overweight Obese | WHO criteria: Underweight Normal Overweight Obese | ORs |

| Author, publication year, country | Title | Data source | Study design | Sample size | Parents age (years, mean(sd)) | Offspring age (years,mean(sd)) | Assessment of anthropometrics | Parental variable(s) | Offspring variable(s) | Reported measurement of association |
|---|---|---|---|---|---|---|---|---|---|---|
| | | clinic in the capital | | | | | | | | |
| Hochner et al.[28] 2012 Israel | Associations of maternal prepregnancy body mass index and gestational weight gain with adult offspring cardiometabolic risk factors: the Jerusalem Perinatal Family Follow-up Study | The Jerusalem Perinatal Study (JPS) population-based cohort | Cohort study | 1,256 | M: 28.38 (5.47) | 32 | M: Self-reported<br>C: Measured | mppBMI(quartile)<br>1:<21.0kg/ $m^2$<br>2:21.0-23.8kg/ $m^2$<br>3:23.9-26.4kg/ $m^2$<br>4:>26.4kg/$m^2$<br>&continuous | BMI Continuous | β (per kg/$m^2$) |
| Johnson et al.[29] 2012 UK | Intergenerational change and familial aggregation of body mass index | The Midspan Renfrew and Paisley Study, and Midspan Family Study | Cohort study | 3,729 | F: 26.0 (3.3)<br>M: 25.9 (4.3) | S: 26.5 (4.0)<br>D: 25.9 (5.0) | M: Measured<br>F: Measured<br>C: Measured | WHO criteria:<br>Underweight<br>Normal<br>Overweight<br>Obese | BMI continuous | Standarized β (per SD) |
| Murrin et al.[30] 2012 Ireland* | Body mass index and height over three | The Lifeways study | Cohort study | 529 | MGM: 60.50 (8.54) | M:30.85 (5.70)<br>F:34.43 (5.65) | MGM, MGF,PGM, PGF: Self-reported | WHO criteria:<br>Underweight<br>Normal<br>Overweight | WHO criteria:<br>Underweight<br>Normal<br>Overweight | correlation coefficient |

| Author, publication year, country | Title | Data source | Study design | Sample size | Parents age (years, mean(sd)) | Offspring age (years,mean(sd)) | Assessment of anthropometrics | Parental variable(s) | Offspring variable(s) | Reported measure-ment of association |
|---|---|---|---|---|---|---|---|---|---|---|
| | generations: evidence from the Lifeways cross-generational cohort study | | | | MGF:63.33 (9.09)<br>PGM:62.10 (8.74)<br>PGF: 62.85 (10.10) | | M: Self-reported<br>F: Self-reported | Obese &BMI continuous | Obese &BMI continuous | |
| Wang et al.[31] 2012 China | Epidemiology of general obesity, abdominal obesity and related risk factors in urban adults from 33 communities of Northeast China: the CHPSNE study | The CHPSNE study(Control Hypertension and Other Risk Factors to Prevent Stroke with Nutrition Education in Urban Area of Northeast China) | Cross-sectional study | 25,196 | NA | 41.7 (14.4) | M: Interview or clinical measure<br>F: Interview or clinical measure<br>C: Interview or clinical measure | WHO Chinese criteria:<br>Underweight<br>Normal<br>Overweight (⩾25 and <27.5 kg/m$^2$)<br>Obese (⩾27.5 kg/m$^2$) | WHO Chinese criteria:<br>Underweight<br>Normal<br>Overweight (⩾25 and <27.5)<br>Obese (⩾27.5 kg/m$^2$)<br>And WHO criteria for Europids (comparable to other studies) | ORs |
| Hu et al.[32] 2013 China | Familial correlation and aggregation of body mass index and blood pressure in Chinese Han population | The China National Nutrition and Health Survey 2002 | Cross-sectional study | 19,107 | F: 52.0 (6.2 )<br>M: 49.8 (5.8) | S:24.8 (5.0)<br>D: 23.0 (4.7) | M:Measured<br>F: Measured<br>C: Measured | Overweight ⩾25 and <30 kg/m$^2$<br>Obese ⩾30 kg/m$^2$<br>&continuous | Overweight ⩾25 and <30 kg/m$^2$<br>Obese ⩾30 kg/m$^2$<br>&continuous | Standarized β(per SD) |

| Author, publication year, country | Title | Data source | Study design | Sample size | Parents age (years, mean(sd)) | Offspring age (years,mean(sd)) | Assessment of anthropometrics | Parental variable(s) | Offspring variable(s) | Reported measurement of association |
|---|---|---|---|---|---|---|---|---|---|---|
| Kelly et al.[33] 2014 Ireland* | Body mass index is associated with the maternal lines but height is heritable across family lines in the Lifeways Cross-Generation Cohort Study | The Lifeways Cross-Generation Cohort Study | Cohort study | 556 families | NA | NA | MGM, MGF, PGM, PGF: Self-reported and measured mixed<br>M: Self-reported<br>F: Self-reported | BMI continuous | BMI continuous | correlation coefficient |
| Vik et al.[34] 2014 Norway | Comparison of father-offspring and mother-offspring associations of cardiovascular risk factors: family linkage within the population-based HUNT Study, Norway | The HUNT study | Cross-sectional study | 36,528 (father-mother–offspring trios) | M: 59.4 (12.5)<br>F: 61.7 (12.4) | 35.6(10.6) | M: Measured<br>F: Measured<br>C: Measured | BMI continuous | BMI continuous | β (Per kg/$m^2$) |
| Cho et al.[35] 2015 USA | Comparisons of chewing rhythm, craniomandib | Sample from local high schools | Cross-sectional study | 32 mother–daug | 49.9 (5.5) | 17.3 (2.2) | NA | BMI continuous | BMI continuous | correlation coefficient |

| Author, publication year, country | Title | Data source | Study design | Sample size | Parents age (years, mean(sd)) | Offspring age (years,mean(sd)) | Assessment of anthropometrics | Parental variable(s) | Offspring variable(s) | Reported measure-ment of association |
|---|---|---|---|---|---|---|---|---|---|---|
| | ular morphology, body mass and height between mothers and their biological daughters. Archives of Oral Biology | and the University of Michigan School of Dentistry | | hter pairs | | | | | | |
| Derraik et al.[36] 2015 Sweden | Obesity rates in two generations of Swedish women entering pregnancy, and associated obesity risk among adult daughters. | Retrospective cohort from the Swedish Birth Register | Cohort study | 26,561 pairs | 26.2 (5.0) | 22.4 (2.3) | M: Weight was measured and current height was self-reported or measured<br>D: same | WHO criteria:<br>Underweight<br>Normal<br>Overweight<br>Obese | WHO criteria:<br>Underweight<br>Normal<br>Overweight<br>Obese | ORs |
| Han et al.[37] 2015 Scottland | Contributions of maternal and paternal adiposity and smoking to adult offspring adiposity and cardiovascular risk: the Midspan Family Study | The Midspan Family Study | Cross-sectional study | 2,230 | F: 54.9(5.0)<br>M: 52.8 (4.9) | Range: 30–59<br>S:44.8 (6.3)<br>D:45.2 (6.1) | M: Measured<br>F: Measured<br>C: Measured | WHO criteria:<br>Underweight<br>Normal<br>Overweight<br>Obese<br>&continuous | WHO criteria:<br>Underweight<br>Normal<br>Overweight<br>Obese<br>&continuous | β (per kg/$m^2$)<br>ORs |

| Author, publication year, country | Title | Data source | Study design | Sample size | Parents age (years, mean(sd)) | Offspring age (years,mean(sd)) | Assessment of anthropometrics | Parental variable(s) | Offspring variable(s) | Reported measure-ment of association |
|---|---|---|---|---|---|---|---|---|---|---|
| Eriksson et al.[38] 2015 Finland | Maternal weight in pregnancy and offspring body composition in late adulthood: findings from the Helsinki Birth Cohort Study (HBCS) | The Helsinki Birth Cohort Study (HBCS) | Longitudinal study | 2,003 | 28.6 (5.5) | Mean age: 62 y | M: Measured<br>C: Measured | Quartile<br>⩽24.6 kg/m$^2$<br>-26.3 kg/m$^2$<br>-28.1 kg/m$^2$<br>>28.1 kg/m$^2$ | FM%<br>LM% | β (per kg/m$^2$) |
| Alati et al.[39] 2016 Australia | Generational increase in obesity among young women: a prospective analysis of mother–daughter dyad | The Mater University Study of Pregnancy (MUSP) | Cohort study | 953 pairs | 18-25 | 21 | M: Measured<br>C: Measured | WHO criteria:<br>Underweight<br>Normal<br>Overweight<br>Obese<br>HW: BMI<25 kg/m$^2$<br>OW/OB: BMI ⩾25 kg/m$^2$ | WHO criteria:<br>Underweight<br>Normal<br>Overweight<br>Obese | ORs |
| Zalbahar et al.[40] 2016 Australia | Parental pre-pregnancy BMI influences on offspring BMI and waist circumference at 21 years | The Mater-University of Queensland Study of Pregnancy (MUSP) cohort | Cohort study | 2,229 pairs | NA | 21 | M: Measured<br>F: Reported by the mother<br>C: Measured | WHO criteria, then collapsed to two groups: Normal weight(<25 kg/m$^2$) or OW/OB(⩾25 kg/m$^2$) | BMI continuous, WHO criteria, then collapsed to two groups: Normal weight(<25 kg/m$^2$) or | β (per kg/m$^2$)<br>ORs |

| Author, publication year, country | Title | Data source | Study design | Sample size | Parents age (years, mean(sd)) | Offspring age (years,mean(sd)) | Assessment of anthropometrics | Parental variable(s) | Offspring variable(s) | Reported measure-ment of association |
|---|---|---|---|---|---|---|---|---|---|---|
| | | | | | | | | | OW/OB(⩾25 kg/m²) | |
| Rath et al.[41] 2016 Australia | Parental pre-pregnancy BMI is a dominant early-life risk factor influencing BMI of offspring in adulthood | The Western Australian Pregnancy Cohort (Raine) Study | Cohort study | 1,355 | NA | Birth to 22 | M: Measured<br>F: Measured<br>C: Measured | Overweight was defined as BMI ⩾25 kg/m², obesity as BMI ⩾30 kg/m² | 2000 Center for Disease Control and Prevention growth charts Overweight was defined as BMI z-score ⩾85th centile, obesity as BMI z-score ⩾95th centile | ORs |
| Swanton et al.[42] 2017 USA | Body mass index associations between mother and offspring from birth to age 18: the Fels Longitudinal Study | The Fels Longitudinal Study | Cohort study | 427 | M: 30-40 | Birth to 18 | M: Measured<br>C: Measured | BMI continuous | BMI continuous | correlation coefficient β (per sd) |
| Rodrigo M. Carrillo-Larco et al.[43] 2018 Peru | Parental body mass index and blood pressure are associated with higher body mass | Population-based implementation study | Cross-sectional study | 955 | F: 59 (11.6)<br>M: 54 (11.8) | 29 (9.5) | M: Measured<br>F: Measured<br>C: Measured | BMI continuous | BMI continuous | β (per kg/m²) |

| Author, publication year, country | Title | Data source | Study design | Sample size | Parents age (years, mean(sd)) | Offspring age (years,mean(sd)) | Assessment of anthropometrics | Parental variable(s) | Offspring variable(s) | Reported measure-ment of association |
|---|---|---|---|---|---|---|---|---|---|---|
| | index and blood pressure in their adult offspring: a cross-sectional study in a resource-limited setting in northern Peru | | | | | | | | | |
| Kaseva et al.[44] 2018 Finland | Pre-pregnancy overweight or obesity and gestational diabetes as predictors of body composition in offspring twenty years later: evidence from two birth cohort studies | The ESTER Maternal Pregnancy Disorders Study and the Arvo Ylppö Longitudinal Study (AYLS) | Cohort study | 891 | At antenatal visit | 24.1 (1.4) | M: healthcare records & questionnaires C: NA | normoglycemic mothers with pre-pregnancy overweight or obesity; mothers with GDM at any level of maternal BMI; offspring of mothers with pre-pregnancy BMI <25 kg/m$^2$ and no GDM | BMI continuous | RD |
| Schoppa et al.[45] 2018 Finland | Association of Maternal Prepregnancy Weight with Offspring | The Framingham Heart Study | Cohort study | 863 | NA | 33 (10) | M: Self-reported or measured C: Measured | Normal weight (BMI < 25 kg/m$^2$) or overweight/ob | Overweight was defined as BMI 25 to 29.9 kg/m$^2$, and obesity | ORs Exposure is owob, outcome is continuous |

| Author, publication year, country | Title | Data source | Study design | Sample size | Parents age (years, mean(sd)) | Offspring age (years,mean(sd)) | Assessment of anthropometrics | Parental variable(s) | Offspring variable(s) | Reported measure-ment of association |
|---|---|---|---|---|---|---|---|---|---|---|
| | Adiposity Throughout Adulthood over 37 Years of Follow-up | Offspring cohort | | | | | | esity (BMI ⩾ 25 kg/m$^2$) | was defined as BMI ⩾ 30 kg/m$^2$ | |
| Chaparro et al.[46] 2017 Sweden | Maternal pre-pregnancy BMI and offspring body composition in young adulthood: the modifying role of offspring sex and birth order | The Uppsala Family Study (UFS) | Longitudinal study | 226 (113 sibling pairs) | 27-29 | 20.2 (15.3-24.6) | M: Self-reported<br>C: Measured | Maternal pre-pregnancy overweight/obesity ( ⩾ 25 kg/m$^2$) | FM%<br>LM% | β (per kg/m$^2$) |

Maternal (M); Paternal(F); Children (C); Daughter(D); Son(S); MGM, Maternal Grandmother; MGF, Maternal Grandfather; PGM, Paternal Grandmother; PGF, Paternal Grandfather; OWOB, Overweight or obesity; Offspring OGDM, mother with gestational diabetes; mppBMI, Maternal pre-pregnancy BMI; NA, not available; FM, fat mass; LM, lean mass

*three-generation study, we extracted data for the first two generations (grandparents and parents);

**Table S2. Descriptions of studies reporting correlation coefficient**

| Study (Author, year) | Title | Methods | Adjusted covariates | Family relationship | r | n |
|---|---|---|---|---|---|---|
| Khoury et al.[1] 1983 | Parent-offspring and sibling body mass index associations during and after sharing of common household environments: the princeton school district family study | correlation | NA | father-son | 0.104 | 34 |
| | | | | father-daughter | 0.132 | 36 |
| | | | | mother-son | 0.167 | 43 |
| | | | | mother-daughter | -0.076 | 49 |
| Friedlander et al.[2] 1988 | Familial aggregation of body mass index in ethnically diverse families in Jerusalem. The Jerusalem Lipid Research Clinic | Interclass correlation | NA | father-son | 0.272 | 1770 |
| | | | | mother-son | 0.205 | 1960 |
| | | | | mother-daughter | 0.169 | 1374 |
| | | | | father-daughter | 0.206 | 1243 |
| | | | | parent-offspring | 0.217 | 6347 |
| Rotimi et al.[4] 1995 | Familial resemblance for anthropometric measurements and relative fat distribution among African Americans | Intraclass correlation | age | mother-son | 0.26 | 65 |
| | | | | mother-daughter | 0.33 | 86 |
| Lake et al.[5] 1997 | Child to adult body mass index in the 1958 British birth cohort associations with parental obesity | Partial correlation (23y) | NA | father-son | 0.21 | 4924 |
| | | | | father-daughter | 0.17 | 4943 |
| | | | | mother-son | 0.24 | 5030 |
| | | | | mother-daughter | 0.25 | 5087 |
| Lake et al.[5] 1997 ¤ | Child to adult body mass index in the 1958 British birth cohort associations with parental obesity | Partial correlation (33y) | NA | father-son | 0.20 | 4403 |
| | | | | father-daughter | 0.15 | 4491 |
| | | | | mother-son | 0.21 | 4496 |
| | | | | mother-daughter | 0.24 | 4644 |
| Williams et al.[8] 2001 | Overweight at age 21 the association with body mass index in childhood and adolescence and parents' body mass index. A | NA | NA | father-son | 0.30 | 482 |
| | | | | mother-son | 0.24 | 482 |
| | | | | mother-daughter | 0.23 | 442 |
| | | | | father-daughter | 0.23 | 442 |

| Study (Author, year) | Title | Methods | Adjusted covariates | Family relationship | r | n |
|---|---|---|---|---|---|---|
| | cohort study of New Zealanders born in 1972–1973 | | | | | |
| Magnusson et al.[10] 2002 | Familial resemblance of body mass index and familial risk of high and low body mass index. A study of young men in Sweden | Pearson correlation | NA | father-son* | 0.280 | 22517 |
| | | | | Quasi father-son** | 0.060 | 1576 |
| Salces et al.[11] 2002 | Family resemblance for anthropometric traits II. Assessment of maternal occupational and age effects | NA | NA | father-offsping^ | 0.601 | 26 |
| | | | | mother-offspring^ | 0.241 | 54 |
| | | | | father-offsping^^ | 0.029 | 9 |
| | | | | mother-offspring^^ | 0.142 | 24 |
| Mirmiran et al.[12] 2002 | Familial clustering of obesity and the role of nutrition: Tehran Lipid and Glucose Study | Bivariate familial correlation | NA | father-son | 0.19 | 27 |
| | | | | mother-son | 0.06 | 27 |
| | | | | mother-daughter | 0.29 | 45 |
| | | | | father-daughter | 0.31 | 45 |
| Wu et al.[14] 2003 | Familial resemblance of adiposity-related parameters Results from a health check-up population in Taiwan | Pearson correlation | NA | father-son | 0.19 | 431 |
| | | | | mother-son | 0.15 | 431 |
| | | | | mother-daughter | 0.14 | 431 |
| | | | | father-daughter | 0.12 | 431 |
| | | Partial correlation | age | father-son | 0.22 | 431 |
| | | | | mother-son | 0.14 | 431 |
| | | | | mother-daughter | 0.13 | 431 |
| | | | | father-daughter | 0.15 | 431 |
| Magarey et al.[15] 2003 | Predicting obesity in early adulthood from childhood and parental obesity | Pearson’s correlation | NA | father-son | 0.20 | 86 |
| | | | | mother-son | 0.35 | 96 |
| | | | | mother-daughter | 0.32 | 87 |

| Study (Author, year) | Title | Methods | Adjusted covariates | Family relationship | r | n |
|---|---|---|---|---|---|---|
| | | | | father-daughter | 0.51 | 70 |
| Classen et al.[24] 2010 | Measures of the intergenerational transmission of body mass index between mothers and their children in the United States, 1981–2004 | correlation | NA | mother-offspring | 0.35 | 4748 |
| | | | | mother-daughter | 0.379 | 2348 |
| | | | | mother-son | 0.319 | 2400 |
| Cooper et al.[25] 2010 | Associations between parental and offspring adiposity up to midlife the contribution of adult lifestyle factors in the 1958 British Birth Cohort Study | correlation | NA | father-son | 0.21 | 3470 |
| | | | | father-daughter | 0.14 | 3503 |
| | | | | mother-son | 0.21 | 3537 |
| | | | | mother-daughter | 0.23 | 3620 |
| Reynold et al[26]. 2010 | Maternal BMI, parity, and pregnancy weight gain: influences on offspring adiposity in young adulthood | Pearson's correlation | NA | mother-offspring | 0.21 | 276 |
| | | | age, sex, smoking status, social class, and activity level | mother-offspring | 0.35 | 276 |
| Murrin et al.[30] 2012 | Body mass index and height over three generations evidence from the Lifeways cross-generational cohort study | Univariate correlation | NA | father-son | -0.059 | 25 |
| | | | | mother-son | 0.085 | 42 |
| | | | | mother-daughter | 0.269 | 128 |
| | | | | father-daughter | 0.302 | 171 |
| Kelly et al.[33] 2014 | Body mass index is associated with the maternal lines but height is heritable across family lines in the Lifeways Cross-Generation Cohort Study | Mixed model | age | father-son | 0.072 | 196 |
| | | | | mother-son | 0.255 | 125 |
| | | | | mother-daughter | 0.245 | 321 |
| | | | | father-daughter | 0.070 | 201 |
| Swanton et al.[42] 2017 | Body mass index associations between mother and offspring from birth to age 18: the Fels Longitudinal Study | Spearman correlation | NA | mother-son | 0.264 | 74 |
| | | | | mother-daughter | 0.269 | 66 |

| Study (Author, year) | Title | Methods | Adjusted covariates | Family relationship | r | n |
|---|---|---|---|---|---|---|
| **Sensitivity analyses or narrative** | | | | | | |
| Cho et al.[35] 2015 | Comparisons of chewing rhythm, craniomandibular morphology, body mass and height between mothers and their biological daughters | Pearson's correlation | NA | mother-daughter | 0.340 | 32 |
| Sørensen et al.[3] 1992 | Correlations of body mass index of adult adoptees and their biological and adoptive relatives | correlation | NA | Adoptee-biological mother | 0.15 | 540 |
| | | | | Adoptee-biological father | 0.11 | 540 |
| | | | | Adoptee-biological sibling | 0.23 | 540 |
| Kazumi et al.[16] 2005 | Associations of middle-aged mother's but not father's body mass index with 18-year-old son's waist circumferences, birth weight, and serum hepatic enzyme levels | Partial correlation | birth weight, son's BMI | father-son | -0.08 | 139 |
| | | | | mother-son | 0.37 | 136 |

NA, not available; r, correlation coefficient
¤only 33 years old was included in the meta-analyses
^Housewife mothers, ^^working mothers

**Table S3. Descriptions of studies reporting mean difference (MD) or standardized mean difference (SMD)**

| Study | Title | Methods | Adjusted Covariates | Exposure unit | Family relationship | MD or SMD | 95% LL | 95%UL | n |
|---|---|---|---|---|---|---|---|---|---|
| Burke et al.[7] 2001 | Family lifestyle and parental body mass index as predictors of body mass index in Australian children: a longitudinal study | Multivariate regression | 'unsafe' drinking, physical fitness, fat consumption, smoking in offspring, and parental education | $kg/m^2$ | father-son | 0.23 | 0.01 | 0.45 | 104 |
| | | | | | father-daughter | 0.44 | 0.24 | 0.64 | 115 |
| | | | | | mother-son | 0.17 | 0.04 | 0.29 | 104 |
| | | | | | mother-daughter | 0.24 | 0.11 | 0.37 | 115 |
| Laitinen et al.[9] 2001 | Family social class, maternal body mass index, childhood body mass index, and age at menarche as predictors of adult obesity | Linear regression | maternal age; social class | $kg/m^2$ | mother-son | 0.21 | 0.16 | 0.26 | 2876 |
| | | | | | mother-daughter | 0.37 | 0.32 | 0.42 | 3404 |
| Kivimäki et al.[18] 2007 | Substantial intergenerational increases in body mass index are not explained by the fetal overnutrition hypothesis: the Cardiovascular Risk in Young Finns Study | Linear regression | age, sex, maternal age, paternal age, maternal BMI, paternal BMI adjusted simultaneously | $kg/m^2$ | mother-offspring | 0.31 | 0.26 | 0.36 | 1918 |
| | | | | | father-offspring | 0.29 | 0.23 | 0.35 | 1918 |
| | | | age, sex | $kg/m^2$ | mother-offspring | 0.32 | 0.27 | 0.37 | 1918 |
| | | | | | father-offspring | 0.34 | 0.28 | 0.41 | 1918 |
| Abu-Rmeileh NM et al.[19] 2008 | Contribution of Midparental BM I and other determinants of ob esity in adult offspring. | Linear regression | sex, age, social class, smoking habit, physical activity, and reported dietary intake | $kg/m^2$ | mid-parental BMI with offspring BMI | 0.51 | 0.41 | 0.62 | 2162 |
| Koupil et al.[20] 2008 | Social and early-life determinants of overweight and obesity in 18-year-old Swedish men | Linear regression | age, mother'age, parity, mother's education, smoking | $kg/m^2$ | mother-son | 0.39 | 0.30 | 0.47 | 1103 |
| | Maternal Anthropometry Is Associated with the Body | | Unadjusted model | $kg/m^2$ | mother-son | 0.70 | 0.47 | 0.94 | 1076 |
| | | | | | mother-daughter | 0.94 | 0.66 | 1.21 | 1112 |

| Study | Title | Methods | Adjusted Covariates | Exposure unit | Family relationship | MD or SMD | 95% LL | 95%UL | n |
|---|---|---|---|---|---|---|---|---|---|
| Tequeanes et al.[21] 2009 | Mass Index and Waist: Height Ratio of Offspring at 23 Years of Age | Multiple linear regressions | maternal age, maternal smoking, maternal parity, maternal education, family income, and skin color * | $kg/m^2$ | mother-son | 0.77 | 0.53 | 1.01 | 1076 |
| | | | | | mother-daughter | 0.94 | 0.66 | 1.22 | 1112 |
| Cooper et al.[25] 2010 | Associations between parental and offspring adiposity up to midlife: the contribution of adult lifestyle factors in the 1958 British Birth Cohort Study | Multiple linear regression | parental age, lifestyle factors, and markers of socioeconomic position | $kg/m^2$ | father-son | 0.27 | 0.23 | 0.31 | 4651 |
| | | | | | father-daughter | 0.22 | 0.17 | 0.28 | 4695 |
| | | | | | mother-son | 0.21 | 0.18 | 0.25 | 4651 |
| | | | | | mother-daughter | 0.29 | 0.25 | 0.34 | 4695 |
| Hochner et al.[28] 2012 | Associations of Maternal Prepregnancy Body Mass Index and Gestational Weight Gain With Adult Offspring Cardiometabolic Risk Factors | Linear regression | ethnicity, sex, maternal and offspring characteristics at the time of birth (ie, parity, mother's age, maternal smoking, socioeconomic status, mother's years of education, maternal medical condition, birth weight, and gestational week) and offspring characteristics at 32 years of age | $kg/m^2$ | mother-offspring* | 0.48 | 0.38 | 0.59 | 1248 |
| Han et al.[37] 2015 | Contributions of maternal and paternal adiposity and smoking to adult offspring adiposity and cardiovascular risk: the Midspan Family Study | Linear Mixed effect models | family clustering, parental and offspring age, smoking and social class | $kg/m^2$ | father-son | 0.35 | 0.27 | 0.42 | 1025 |
| | | | | | father-daughter | 0.29 | 0.21 | 0.38 | 1283 |
| | | | | | mother-son | 0.26 | 0.20 | 0.32 | 1025 |
| | | | | | mother-daughter | 0.33 | 0.27 | 0.40 | 1283 |
| | | | Mutually adjust for parental BMI | $kg/m^2$ | father-son | 0.30 | 0.23 | 0.38 | 1025 |
| | | | | | father-daughter | 0.23 | 0.15 | 0.32 | 1283 |

| Study | Title | Methods | Adjusted Covariates | Exposure unit | Family relationship | MD or SMD | 95% LL | 95%UL | n |
|---|---|---|---|---|---|---|---|---|---|
| | | | | | mother-son | 0.22 | 0.16 | 0.28 | 1025 |
| | | | | | mother-daughter | 0.33 | 0.24 | 0.37 | 1283 |
| Zalbahar et al.[40] 2016 | Parental pre-pregnancy BMI influences on offspring BMI and waist circumference at 21 years | Multiple linear regression | Unadjusted model | $kg/m^2$ | father-offspring | 0.36 | 0.30 | 0.42 | 2229 |
| | | | | | mother-offspring | 0.35 | 0.31 | 0.40 | 2229 |
| | | | Other parent's BMI, offspring sex, maternal factor, maternal and paternal education, annual family income (model2) | $kg/m^2$ | father-offspring | 0.36 | 0.29 | 0.42 | 2229 |
| | | | | | mother-offspring | 0.38 | 0.32 | 0.43 | 2229 |
| | | | Unadjusted model | $kg/m^2$ | father-son | 0.30 | 0.23 | 0.38 | 1114 |
| | | | | | father-daughter | 0.41 | 0.32 | 0.50 | 1115 |
| | | | | | mother-son | 0.25 | 0.19 | 0.32 | 1114 |
| | | | | | mother-daughter | 0.47 | 0.39 | 0.54 | 1115 |
| | | | Other parent's BMI, offspring sex, maternal factor, maternal and paternal education, annual family income (model2) | $kg/m^2$ | father-son | 0.31 | 0.23 | 0.39 | 1114 |
| | | | | | father-daughter | 0.39 | 0.29 | 0.49 | 1115 |
| | | | | | mother-son | 0.30 | 0.22 | 0.37 | 1114 |
| | | | | | mother-daughter | 0.44 | 0.35 | 0.52 | 1115 |
| Rodrigo M. Carrillo-Larco et al.[43] 2018 | Parental body mass index and blood pressure are associated with higher body mass index and blood pressure in their adult offspring: a cross-sectional study in a resource-limited setting in northern Peru | Mixed-effects linear regression | Adjusted by village, age, educational level, physical activity of the offspring and wealth index of the family. | $kg/m^2$ | father-son | 0.26 | 0.14 | 0.38 | 253 |
| | | | | | father-daughter | 0.25 | 0.13 | 0.37 | 185 |
| | | | | | mother-son | 0.20 | 0.10 | 0.31 | 253 |
| | | | | | mother-daughter | 0.11 | −0.00 | 0.22 | 185 |
| | | | Unadjusted model | $kg/m^2$ | father-son | 0.20 | 0.08 | 0.32 | 253 |
| | | | | | father-daughter | 0.19 | 0.07 | 0.32 | 185 |
| | | | | | mother-son | 0.20 | 0.10 | 0.30 | 253 |
| | | | | | mother-daughter | 0.10 | -0.02 | 0.21 | 185 |

| Study | Title | Methods | Adjusted Covariates | Exposure unit | Family relationship | MD or SMD | 95% LL | 95%UL | n |
|---|---|---|---|---|---|---|---|---|---|
| Kvaavik et al.[13] 2003 | Predictors and Tracking of Body Mass Index From Adolescence Into Adulthood | Linear regression | father's education, leisure time physical activity, fitness, smoking, baseline BMI, sex, own education, leisure time physical activity, smoking at FU | SD | mother-offspring | 0.07 | -0.01 | 0.16 | 233 |
| | | | | | father-offsping | 0.12 | 0.03 | 0.21 | 249 |
| Hu et al.[32] 2008 | Familial correlation and aggregation of body mass index and blood pressure in Chinese Han population | Multilevel linear regression | children's age, both childrens' and parents' education level and occupation | SD | father-son | 0.27 | 0.23 | 0.30 | 4132 |
| | | | | | mother-son | 0.25 | 0.21 | 0.27 | 4132 |
| | | | | | father-daughter | 0.23 | 0.18 | 0.27 | 2237 |
| | | | | | mother-daughter | 0.21 | 0.18 | 0.23 | 2237 |
| Johnson et al.[29] 2012 | Intergenerational change and familial aggregation of body mass index | Multilevel linear regression | Age, marital status, number of children, smoking status and social class | SD | father-son | 0.27 | 0.22 | 0.33 | 1023 |
| | | | | | father-daughter | 0.19 | 0.14 | 0.25 | 1263 |
| | | | | | mother-son | 0.28 | 0.22 | 0.33 | 1023 |
| | | | | | mother-daughter | 0.31 | 0.27 | 0.36 | 1263 |
| | | | | | mother-offspring | 0.30 | 0.26 | 0.34 | 1443 |
| | | | | | father-offspring | 0.23 | 0.19 | 0.27 | 1443 |
| Swanton et al.[42] 2017 | Body mass index associations between mother and offspring from birth to age 18: the Fels Longitudinal Study | Multiple linear regression | Maternal birth year | SD | mother-son | 0.27 | -0.00 | 0.54 | 74 |
| | | | | | mother-daughter | 0.46 | -0.00 | 0.92 | 66 |
| | | | Decade of birth, parity | SD | mother-son | 0.28 | -0.00 | 0.56 | 156 |
| | | | | | mother-daughter | 0.30 | -0.00 | 0.60 | 133 |
| Kaseva et al.[44] 2018 | Pre-pregnancy overweight or obesity and gestational diabetes as predictors of body composition in offspring twenty years later: evidence from two birth cohort studies | Linear regression | age, cohort, gestational age, birth weight SD score, maternal hypertension or preeclampsia during pregnancy, maternal smoking during pregnancy and parental educational attainment | OWOB Versus Normal | mother-son | 1.64 | 0.57 | 2.72 | 335 |
| | | | | | mother-daughter | 1.41 | 0.20 | 2.63 | 365 |
| | | | Age, cohort | | mother-son | 2.35 | 1.34 | 3.36 | 335 |

| Study | Title | Methods | Adjusted Covariates | Exposure unit | Family relationship | MD or SMD | 95% LL | 95%UL | n |
|---|---|---|---|---|---|---|---|---|---|
| | | | | Owob vs normal | mother-daughter | 1.67 | 0.56 | 2.78 | 365 |
| Vik et al.[34] 2014 ∅ | Comparison of father-offspring and mother-offspring associations of cardiovascular risk factors family linkage within the population-based HUNT Study, Norway | Linear regression | Age, sex | kg/m$^2$ | father-offspring | *0.22* | *0.20* | *0.23* | 36528 |
| | | | | | mother-offspring | 0.17 | 0.16 | 0.18 | 36528 |
| | | | | SD | father-offspring | *0.19* | *0.18* | *0.20* | 36528 |
| | | | | | mother-offspring | 0.20 | 0.18 | 0.21 | 36528 |
| Eriksson et al.[38] 2015 ∅ | Maternal weight in pregnancy and offspring body composition in late adulthood: findings from the Helsinki Birth Cohort Study (HBCS) | Multiple linear regression | Age and sex | kg/m$^2$ | Mother-offspring | 4.9 | 2.7 | 7.0 | 1650 |
| | | | Age | | Mother-son | 6.9 | 4.0 | 9.8 | 809 |
| | | | | | Mother-daughter | 2.6 | -0.6 | 5.8 | 809 |
| Chaparro et al.[46] 2017 ∅ | Maternal pre-pregnancy BMI and offspring body composition in young adulthood: the modifying role of offspring sex and birth order | Multivariable linear regression | Age, height, mother's age at birth, and mother's education | SD | mother-son | 0.59 | 0.27-1.44 | 226 | 226 |
| | | | | | mother-daughter | 0.97 | 0.14-1.80 | 226 | 226 |
| Schoppa et al.[45] 2019 ∅ | Association of Maternal Prepregnancy Weight with Offspring Adiposity Throughout Adulthood over 37 Years of Follow-up | Linear regression | Age, sex | Owob vs normal | mother-offspring | 1.4 | 0.62 | 2.18 | 863 |
| | | | Age, sex, BMI GRS | Owob vs normal | mother-offspring | 1.6 | 0.62 | 2.58 | 766 |

LL, lower limit; UL, upper limit; MD, mean difference; SMD, standardized mean difference; OWOB, overweight or obese

∅ not included in meta-analyses

**Table S4. Descriptions of studies reporting odds ratios (ORs) or risk ratios (RRs)**

| Author | Title | Exposure Criteria | Outcome Criteria | Family relationship | Comparison group vs reference group | ORs (95%CI) | | | Adjustments |
|---|---|---|---|---|---|---|---|---|---|
| | | | | | | Over-weight | Obese | OWOB | |
| Lake et al.[5] 1997 | Child to adult body mass index in the 1958 British birth cohort associations with parental obesity | Underweight (<20 kg/m$^2$ for father, <18.7 for mother); Normal weight(20-24.9 kg/m$^2$ for father, 18.7-23.7 for mother); Overweight( 25-27.7 kg/m$^2$ for father, 23.7-27.6 for mother); Obesity(>27. 7 kg/m$^2$ for father, >27.6 for mother) | at or above 85th percentile for their age and sex are classified as obesity | parent-son | 2 overweight parents vs both normal weight parents | | 3.41 (2.36-4.93) | | Age |
| | | | | parent-son | 1 obese parent vs both normal weight parents | | 3.43 (2.53-4.64) | | |
| | | | | parent-daughter | 1 overweight parent vs both normal weight parents | | 1.47 (1.14-1.88) | | |
| | | | | parent-son | 2 obese parents vs both normal weight parents | | 8.42 (5.47-13.00) | | |
| | | | | parent-daughter | 2 obese parents vs both normal weight parents | | 6.75 (4.57-9.96) | | |
| | | | | parent-son | 1 overweight parent vs both normal weight parents | | 1.99 (1.45-2.73) | | |
| | | | | parent-daughter | 2 overweight parents vs both normal weight parents | | 2.65 (1.95-3.62) | | |
| | | | | parent-daughter | 1 obese parent vs both normal weight parents | | 2.98 (2.35-3.78) | | |
| Al-Isa et al.[6] 1998 | Factors Associated with | both parents obese; father or mother | Normal weight<25; | mother-daughter | obese mother vs both normal weight parents | 2.02 (1.28-3.19) | 4.27 (2.02-9.05) | | number of brothers, having chronic |

| Author | Title | Exposure Criteria | Outcome Criteria | Family relationship | Comparison group vs reference group | ORs (95%CI) | | | Adjustments |
|---|---|---|---|---|---|---|---|---|---|
| | | | | | | Over-weight | Obese | OWOB | |
| | Overweight and Obesity among Kuwaiti College Women | obese; neither parent obese | 25⩽overweight<30; obese>30 kg/$m^2$ | father-daughter | obese father vs both normal weight parents | 1.46 (0.67-3.20) | 1.31 (0.27-6.34) | | diseases, Having Obesity parent, Dieting, Country prefer visiting |
| Burke et al.[7] 2001 | Family lifestyle and parental body mass index as predictors of body mass index in Australian children: a longitudinal stud | Normal weight<25; 25⩽overweight<30; obese>30 kg/$m^2$ | Normal weight<25; 25⩽overweight<30; obese>30 kg/$m^2$ | mother-daughter | obese mother vs normal weight mother | | | 7.88 (1.97-31.43) | 'unsafe' drinking, physical fitness, fat consumption, smoking in offspring, and parental education |
| | | | | father-daughter | obese father vs normal weight father | | | 3.83 (1.01-14.54) | |
| | | | | mother-son | obese mother vs normal weight mother | | | 2.49 (0.69-8.89) | |
| | | | | father-son | obese father vs normal weight father | | | 4.38 (1.16-16.48) | |
| Williams et al.[8] 2001 | Overweight at age 21: the association with body mass index in childhood and adolescence and parents' body mass index. A cohort study of New Zealanders | underweight <18.5; 18.5⩽normal weight<25; 25⩽overweight<30; obese⩾30 kg/$m^2$ | underweight<18.5; 18.5⩽normal weight<25; 25⩽overweight<30; obese⩾30 kg/$m^2$ | father-offspring | overweight father vs normal weight father | | | 1.2 (0.60-2.00) | Sex, and other parent's BMI, children's BMI at 11 years |
| | | | | mother-offspring | obese mother vs normal weight mother | | | 1.9 (1.20-2.50) | |
| | | | | father-offspring | obese father vs normal weight father | | | 1.3 (1.00-1.70) | |
| | | | | mother-offspring | overweight mother vs normal weight mother | | | 1.1 (0.80-1.60) | |

| Author | Title | Exposure Criteria | Outcome Criteria | Family relationship | Comparison group vs reference group | ORs (95%CI) | | | Adjustments |
|---|---|---|---|---|---|---|---|---|---|
| | | | | | | Over-weight | Obese | OWOB | |
| | born in 1972–1973 | | | | | | | | |
| Kvaavik et al.[13] 2003 | Predictors and Tracking of Body Mass Index From Adolescence Into Adulthood | Normal weight<25; 25⩽overweight<30; obese>30 kg/m² | Normal weight<25; 25⩽overweight<30; obese>30 kg/m² | parent-offspring | overweight parent vs both normal weight parent | 0.54(0.89) | 0.61(1.6) | | Adolescent BMI, Tanner stage, sex,the adolescents' ascribed and family characteristics, perceptions of family relationship,adolescents' self-esteem and weight status adolescent behaviors |
| Crossman et al.[17] 2006 | The family environment and American adolescents' risk of obesity as young adults | not obese: <30, obese⩾30 kg/m² | CDC's age- and sex-specific BMI percentiles for children and adolescents | mother-son | obese mother vs normal weight mother | | | 1.33 | the adolescents' ascribed and family characteristics |
| | | | | father-daughter | obese father vs normal weight father | | | 1.58 | |
| | | | | mother-daughter | obese mother vs normal weight mother | | | 1.39 | |
| | | | | father-son | obese father vs normal weight father | | | 1.6 | |
| Al-Isa et al.[27] 2013 | Factors Associated With Overweight and Obesity | Normal weight⩽25; 25<overweight<30; obese>30 kg/m² | Normal weight⩽25; 25<overweight<30; obese>30 kg/m² | parent-son | 2 obese parents vs both normal weight parents | 5.9 (1.4-25.7) | 2.9 (1.14-7.19) | | Offspring age, dental status, chronic disease, number of Obesity brothers, |
| | | | | mother-son | Obese mother vs both normal weight parents | 2 (1.19-3.68) | 2.4 (1.24-4.74) | | |

| Author | Title | Exposure Criteria | Outcome Criteria | Family relationship | Comparison group vs reference group | ORs (95%CI) | | | Adjustments |
|---|---|---|---|---|---|---|---|---|---|
| | | | | | | Over-weight | Obese | OWOB | |
| | Among Kuwaiti Men | | | father-son | Obese father vs both normal weight parents | 1.9 (0.84-4.4) | 1.3 (0.53-3.2) | | number of Obesity relatives, wife's education, last GPA, high school GPA, monthly family income, physical activity, sport, health status, dieting, feeling tired |
| Wang et al.[31] 2012 | Epidemiology of general obesity, abdominal obesity and related risk factors in urban adults from 33 communities of northeast china: the CHPSNE study | history of obesity | Normal weight<25; 25⩽overweight<27.5; obese⩾27.5 kg/m$^2$ | parent-offspring | 1 obese parent vs both normal weight parents | | 2.61 (2.4-2.83) | | age, ethnicity, education, occupation, family income, physical activity, cigarette smoking, alcohol, eat fried foods, diet |
| | | | | parent-son | 1 obese parent vs both normal weight parents | | 3.51 (3.13-3.94) | | |
| | | | | parent-daughter | 1 obese parent vs both normal weight parents | | 2.06 (1.82-2.33) | | |
| Magarey et al.[47] 2003 | Predicting obesity in early adulthood from childhood and parental obesity | Normal weight<25; 25⩽overweight<30; obese>30 kg/m$^2$ | Normal weight<25; 25⩽overweight<30; obese>30 kg/m$^2$ | parent-offspring | 1 owob parent vs normal weight parent | | | 2.1 (1.0–4.5) | NA |
| | | | | parent-offspring | 2 owob parents vs normal weight parent | | | 4.3 (2.1–8.9) | |

| Author | Title | Exposure Criteria | Outcome Criteria | Family relationship | Comparison group vs reference group | ORs (95%CI) | | | Adjustments |
|---|---|---|---|---|---|---|---|---|---|
| | | | | | | Over-weight | Obese | OWOB | |
| Derraik et al.[36] 2015 | Obesity rates in two generations of Swedish women entering pregnancy and associated obesity risk among adult daughters | underweight <18.5; 18.5⩽normal weight<25; 25⩽overweight<30; obese⩾30 kg/m$^2$ | underweight<18.5; 18.5⩽normal weight<25; 25⩽overweight<30; obese⩾30 kg/m$^2$ | mother-daughter | overweight mother vs normal weight mother | 2.44 (2.26, 2.63) | | | Unadjusted |
| | | | | mother-daughter | obese mother vs normal weight mother | 3.71 (3.18, 4.34) | | | |
| | | | | mother-daughter | overweight mother vs normal weight mother | | 2.71 (2.46, 2.98) | | |
| | | | | mother-daughter | obese mother vs normal weight mother | | 4.65 (3.95, 5.48) | | |
| Han et al.[37] 2015 | Contributions of maternal and paternal adiposity and smoking to adult offspring adiposity and cardiovascular risk: the Midspan Family Study | Normal weight<25; 25⩽overweight<30; obese>30 kg/m$^2$ | Normal weight<25; 25⩽overweight<30; obese>30 kg/m$^2$ | parent-offspring | obese parent vs both normal weight parents | 10.4 (4.65-13.29) | | | family clustering, offspring and parental age, smoking status (either current or former smokers) and social class |
| | | | | parent-offspring | obese parent vs both normal weight parents | 1.96 (1.54-2.48) | | | |
| | | | | parent-offspring | obese parent vs both normal weight parents | | 11.77 (7.70-18.00) | | |
| | | | | parent-offspring | obese parent vs both normal weight parents | | 2.14 (1.62-2.81) | | |
| Alati et al.[39] 2016 | Generational increase in obesity among young women: a | underweight <18.5; 18.5⩽normal weight<25; 25⩽overweight<30; | underweight<18.5; 18.5⩽normal weight<25; 25⩽overweight<30; obese⩾30 kg/m$^2$ | mother-daughter | overweight mother vs normal weight mother | 2.54 (1.86, 3.54) | | | age in years, education, number of previous pregnancies, exercise and |
| | | | | father-son | overweight father vs normal weight father | | 7.85 (3.58-22.83) | | |

| Author | Title | Exposure Criteria | Outcome Criteria | Family relationship | Comparison group vs reference group | ORs (95%CI) | | | Adjustments |
|---|---|---|---|---|---|---|---|---|---|
| | | | | | | Over-weight | Obese | OWOB | |
| | prospective analysis of mother–daughter dyads | obese⩾30 kg/m² | | father-daughter | overweight father vs normal weight father | 1.52 (1.16-2.08) | | | television viewing |
| | | | | father-son | overweight father vs normal weight father | 1.91 (1.29-2.93) | | | |
| | | | | mother-son | obese mother vs normal weight mother | | 4.01 (2.36-7.46) | | |
| | | | | mother-son | overweight mother vs normal weight mother | 2.83 (2.03-4.13) | | | |
| | | | | mother-daughter | obese mother vs normal weight mother | | 5.04 (3.03, 8.85) | | |
| | | | | father-daughter | obese father vs normal weight father | | 8.33 (5.10-15.96) | | |
| Zalbahar et al.[40] 2016 | Parental pre-pregnancy BMI influences on offspring BMI and waist circumference at 21 years | underweight <18.5; 18.5⩽normal weight<25; 25⩽overweight<30; obese⩾30 kg/m² | underweight<18.5; 18.5⩽normal weight<25; 25⩽overweight<30; obese⩾30 kg/m² | father-offspring | owob father vs normal weight father | | | 2.39 (1.82-3.14) | other parent's, offspring's sex, gestational weight gain, maternal age at birth, maternal smoking during pregnancy), maternal and paternal education attainment, annual family income |
| | | | | mother-offspring | owob mother vs normal weight mother | | | 2.36 (1.69-3.29) | |
| | | | | parent-offspring | both owob parent vs normal weight parent | | | 5.73 (3.34-9.83) | |

| Author | Title | Exposure Criteria | Outcome Criteria | Family relationship | Comparison group vs reference group | ORs (95%CI) | | | Adjustments |
|---|---|---|---|---|---|---|---|---|---|
| | | | | | | Over-weight | Obese | OWOB | |
| Rath et al.[41] 2016 | Parental pre-pregnancy BMI is a dominant early-life risk factor | Normal weight<25; 25⩽overweight<30; obese>30 kg/m$^2$ | at or above 95th percentile for their age and sex are classified as obesity | father-offspring | overweight father vs normal weight father | | 1.44 (0.92-2.23) | | Sex, maternal education, maternal smoking, maternal anemia and diabetes, cesarean, prematurity, birth weight, first year weight gain, duration of breast-feeding |
| | | | | mother-offspring | obese mother vs normal weight mother | | 6.96 (3.81-12.52) | | |
| | | | | mother-offspring | overweight mother vs normal weight mother | | 4.58 (2.87-7.24) | | |
| | | | | father-offspring | obese father vs normal weight father | | 3.75 (1.80-7.50) | | |
| Stuebe et al.[23] 2009 | Maternal-recalled gestational weight gain, pre-pregnancy body mass index, and obesity in the daughter | BMI cut off: 21, 23, 25, 29 kg/m$^2$ | Normal weight<25; 25⩽overweight<30; obese>30 kg/m$^2$ | mother-daughter | mother BMI=29 vs BMI =21 kg/m2 | | 7.53 (6.00-9.45) | | Unadjusted |
| Koupil et al.[20] 2008 | Social and early-life determinants of overweight and obesity in 18-year-old Swedish men | continuous | Normal weight<25; 25⩽overweight<30; obese>30 kg/m$^2$ | mother-son | per 1 kg/m$^2$ increase in mother's mean BMI | | | 1.23 (1.16-1.30) | age, mother's age, parity and mother's education, smoking |
| Kowalski-Jones et al.[48] 2009 | Are You What Your Mother Weighs? | continuous | at or above 95th percentile for their age and sex | mother-son | per 1 kg/m$^2$ increase in mother's mean BMI | 1.11(1.07–1.16) | | | family income, marriage time, residence, maternal weeks |

| Author | Title | Exposure Criteria | Outcome Criteria | Family relationship | Comparison group vs reference group | ORs (95%CI) | | | Adjustments |
|---|---|---|---|---|---|---|---|---|---|
| | | | | | | Over-weight | Obese | OWOB | |
| | Evaluating the Impact of Maternal Weight Trajectories on Youth Overweight | | are classified as overweight | mother-daughter | per 1 $kg/m^2$ increase in mother's mean BMI | 1.06(1.07–1.15) | | | worked, neighborhood quality |
| Reynolds et al.[26] 2010 | Maternal BMI, Parity, and Pregnancy Weight Gain: Influences on Offspring Adiposity in Young Adulthood | continuous | overweight and obese vs normal | mother-offspring | per 1 SD increase in mother's mean BMI | | | 1.99 | Gender, smoking, maternal age, antenatal weight gain, primiparous |

Note: all studies used logistic regression, except Han et al.[37], which used Generalized estimating equation model.

BMI, body mass index; OR, odds ratio; CI, confidence interval; OWOB, overweight or obese; SD, standard deviation

**Table S5. Pooled standardized mean difference (SMD) between parental and offspring BMI (per SD)**

| Series | Family relationship | Pooled SMD for adjusted models | | | | Pooled SMD for unadjusted models* | | | |
|---|---|---|---|---|---|---|---|---|---|
| | | SMD | 95%CI | $I^2$ (%) | Study number | SMD | 95%CI | $I^2$ (%) | Study number |
| **Level1** | Mother-daughter | 0.25 | 0.21-0.29 | 70.5 | 10 | 0.25 | 0.20-0.30 | 81.4 | 17 |
| | Mother-son | 0.23 | 0.20-0.26 | 61.8 | 11 | 0.23 | 0.20-0.27 | 56.0 | 16 |
| | Father-daughter | 0.21 | 0.16-0.26 | 68.5 | 7 | 0.20 | 0.16-0.24 | 67.1 | 12 |
| | Father-son | 0.25 | 0.21-0.28 | 39.9 | 7 | 0.23 | 0.19-0.27 | 77.7 | 13 |
| **Level2** | Mother-offspring | 0.23 | 0.20-0.26 | 78.9 | 13 | 0.24 | 0.21-0.28 | 84.6 | 18 |
| | Father-offspring | 0.22 | 0.19-0.25 | 68.1 | 9 | 0.21 | 0.18-0.25 | 71.5 | 13 |

SMD, standardized mean difference; SD, standardized deviation; CI, Confidence interval

*include studies reporting correlation coefficient and unadjusted SMD in regression coefficient

$I^2$ = proportion of total variation in effect estimate due to between-study heterogeneity (based on Q)

**Table S6. Pooled mean difference (MD) between parental and offspring BMI (per kg/m$^2$)**

| Series | Family relationship | Pooled MD for adjusted models | | | | Pooled MD for unadjusted models | | | |
|---|---|---|---|---|---|---|---|---|---|
| | | MD | 95%CI | I² (%) | Study number | MD | 95%CI | I² (%) | Study number |
| **Level1** | Mother-daughter | 0.34 | 0.26-0.43 | 87.6 | 7 | 0.48 | 0.13-0.83 | 95.5 | 3 |
| | Mother-son | 0.27 | 0.21-0.34 | 82.4 | 8 | 0.34 | 0.16-0.52 | 86.6 | 3 |
| | Father-daughter | 0.30 | 0.22-0.38 | 67.7 | 5 | 0.30 | 0.09-0.52 | 86.7 | 2 |
| | Father-son | 0.29 | 0.26-0.32 | 3.1 | 5 | 0.26 | 0.17-0.36 | 47.9 | 2 |
| **Level2** | Mother-offspring | 0.34 | 0.28-0.41 | 90.7 | 9 | 0.38 | 0.24-0.51 | 93.7 | 4 |
| | Father-offspring | 0.30 | 0.26-0.35 | 65.0 | 6 | 0.30 | 0.21-0.39 | 80.5 | 3 |

MD, mean difference; CI, Confidence interval

$I^2$ = proportion of total variation in effect estimate due to between-study heterogeneity (based on Q)

**Table S7. Difference of standardized mean difference between maternal and paternal line in adjusted models**

| Author | Diff. Of SMD | 95% CI | | % Weight | Test of overall effect | | |
|---|---|---|---|---|---|---|---|
| | | LL | UL | | p value compare MO and FO | I² (%) | tau² |
| Burke | -0.02 | -0.19 | 0.16 | 3.02 | 0.49 | 46.8 | 0.0011 |
| Cooper | 0.01 | -0.03 | 0.05 | 18.42 | | | |
| Han | 0.03 | -0.04 | 0.10 | 11.51 | | | |
| Hu | -0.03 | -0.06 | 0.01 | 21.15 | | | |
| Johnson | 0.07 | 0.01 | 0.13 | 14.7 | | | |
| Kivimäki | 0.06 | -0.02 | 0.14 | 10.34 | | | |
| Kvaavik | -0.05 | -0.17 | 0.07 | 5.32 | | | |
| Carrillo-Larco | -0.09 | -0.21 | 0.02 | 6.08 | | | |
| Zalbahar | 0.04 | -0.05 | 0.12 | 9.46 | | | |
| Overall, DL | 0.01 | -0.02 | 0.05 | 100 | | | |

SMD, Standardized Mean Difference; MO, mother-offspring; FO, father-offspring; LL, lower limit; UL, upper limit

$I^2$ = proportion of total variation in effect estimate due to between-study heterogeneity (based on Q)

**Table S8. Difference of standardized mean difference between maternal and paternal line in unadjusted models**

| Author | Diff. in SMD | 95% CI | | % Weight | Test of overall effect | | |
|---|---|---|---|---|---|---|---|
| | | LL | UL | | p value compare MO and FO | I² (%) | tau² |
| Cooper | 0.05 | 0.01 | 0.08 | 19.75 | 0.82 | 65.5 | 0.003 |
| Friedlander | -0.06 | -0.11 | -0.01 | 17.52 | | | |
| Kelly | 0.18 | 0.05 | 0.32 | 7.43 | | | |
| Khoury | -0.08 | -0.41 | 0.24 | 1.82 | | | |
| Lake | 0.05 | 0.02 | 0.08 | 20.21 | | | |
| Magarey | -0.01 | -0.23 | 0.20 | 3.66 | | | |
| Mirmiran | -0.06 | -0.40 | 0.28 | 1.65 | | | |
| Murrin | -0.04 | -0.25 | 0.17 | 3.92 | | | |
| Salces | -0.34 | -0.38 | -0.30 | 1.07 | | | |
| William | -0.03 | -0.13 | 0.06 | 11.68 | | | |
| Wu | -0.05 | -0.15 | 0.04 | 11.29 | | | |
| Overall, DL | 0.01 | -0.04 | 0.05 | 100 | | | |

SMD, Standardized Mean Difference; MD, mean difference; MO, mother-offspring; FO, father-offspring; LL, lower limit; UL, upper limit

$I^2$ = proportion of total variation in effect estimate due to between-study heterogeneity (based on Q)

**Table S9. Difference of mean difference between maternal and paternal line**

| Series | Author | Difference of MDs | 95% CI | | % Weight | Test of overall effect | | |
|---|---|---|---|---|---|---|---|---|
| | | | LL | UL | | p value compare MO and FO | $I^2$ (%) | $tau^2$ |
| i.MD adjusted covariates | Burke | -0.14 | -0.32 | 0.03 | 3.66 | 0.29 | 14.1 | 0.0003 |
| | Cooper | -0.01 | -0.06 | 0.03 | 40.18 | | | |
| | Han | -0.03 | -0.10 | 0.04 | 18.49 | | | |
| | Kivimäki | 0.02 | -0.06 | 0.10 | 15.9 | | | |
| | Carrillo-Larco | -0.10 | -0.21 | 0.02 | 8.08 | | | |
| | Zalbahar | 0.02 | -0.07 | 0.11 | 13.69 | | | |
| | Overall, DL | -0.02 | -0.05 | 0.02 | 100 | | | |
| ii.MD without adjusting for covariates | Kivimäki | -0.02 | -0.10 | 0.06 | 39.26 | 0.45 | 0.0 | 0.0000 |
| | Rodrigo M. Carrillo-Larco | -0.04 | -0.15 | 0.08 | 18.16 | | | |
| | Zalbahar | -0.01 | -0.09 | 0.07 | 44.19 | | | |
| | Overall, DL | -0.02 | -0.07 | 0.03 | 100 | | | |

MD, mean difference; CI, Confidence interval; MO, mother-offspring; FO, father-offspring; LL, lower limit; UL, upper limit

$I^2$ = proportion of total variation in effect estimate due to between-study heterogeneity (based on Q)

**Table S10. Summary of studies not included in meta-analyses**

| Study | Title | Parental BMI categories | Offspring outcome | Main finding | Reason not included |
|---|---|---|---|---|---|
| Chaparro et al.[46] 2017 | Maternal pre-pregnancy BMI and offspring body composition in young adulthood: the modifying role of offspring sex and birth order | Maternal pre-pregnancy BMI | % fat mass<br>% lean mass | A higher pre-pregnancy BMI was associated with higher offspring % fat mass and lower offspring % lean mass in late adolescence and young adulthood | Outcome is body fat percentage |
| Eriksson et al.[38] 2015 | Maternal weight in pregnancy and offspring body composition in late adulthood: findings from the Helsinki Birth Cohort Study (HBCS) | Maternal BMI >28.1 kg/$m^2$, Maternal BMI <28.1 kg/$m^2$ | % fat mass<br>% lean mass | Higher maternal BMI was associated with less favorable body composition in the offspring. | Outcome is body fat percentage |
| Vik et al.[34] 2013 | Comparison of father-offspring and mother-offspring associations of cardiovascular risk factors family linkage within the population-based HUNT Study, Norway | Parental BMI as continuous | Offspring BMI continuous | This study found similar father-offspring and mother-offspring associations across all cardiovascular risk factors under study | The study reported residuals from linear regression analysis, and therefore not amenable to inclusion in the meta-analysis |
| Sørensen et al.[3] 1992 | Correlations of body mass index of adult adoptees and their biological and adoptive relatives | BMI continuous | BMI continuous | The study found correlations between adoptees and their biological parents, but no correlation in BMI between the adoptees and their adoptive parents | The adoption study setting is different from other studies |

**Table S11. Subgroup analyses by BMI measurement method**

| Categories | Family relationship | Self-reported | | | | Measured | | | | Overall I² (%) |
|---|---|---|---|---|---|---|---|---|---|---|
| | | SMD | 95%CI | Study numbers | I² (%) | SMD | 95%CI | Study numbers | I² (%) | |
| Offspring's BMI measurement methods | Mother-offspring | 0.14 | 0.01-0.27 | 2 | 87.2 | 0.25 | 0.22-0.28 | 12 | 73.3 | 77.5 |
| | Father-offspring | 0.12 | 0.03-0.21 | 1 | 0.0 | 0.23 | 0.20-0.26 | 8 | 65.0 | 68.1 |
| Parents'BMI measurement methods | Mother-offspring | 0.21 | 0.15-0.27 | 6 | 79.7 | 0.26 | 0.23-0.30 | 7 | 66.5 | 77.5 |
| | Father-offspring | 0.17 | 0.13-0.22 | 3 | 42.7 | 0.25 | 0.23-0.27 | 6 | 0.0 | 68.1 |

SMD, standardized mean difference; CI, confidence interval; NA, not available

$I^2$ = proportion of total variation in effect estimate due to between-study heterogeneity (based on Q)

**Table S12. Subgroup analyses by study design**

| Family relationship | Cohort studies | | | | Cross-sectional studies | | | | Overall $I^2$ (%) |
|---|---|---|---|---|---|---|---|---|---|
| | SMD | 95%CI | Study numbers | $I^2$ (%) | SMD | 95%CI | Study numbers | $I^2$ (%) | |
| Mother- daughter | 0.26 | 0.20-0.32 | 6 | 71.8 | 0.24 | 0.18-0.30 | 4 | 65.3 | 70.5 |
| Mother-son | 0.22 | 0.17-0.26 | 7 | 65.3 | 0.25 | 0.23-0.28 | 4 | 0.00 | 61.80 |
| Father-daughter | 0.17 | 0.11-0.24 | 3 | 69.2 | 0.24 | 0.19-0.31 | 4 | 41.3 | 68.5 |
| Father-son | 0.23 | 0.18-0.28 | 3 | 58.3 | 0.27 | 0.24-0.30 | 4 | 0.00 | 39.9 |
| Mother-offspring | 0.23 | 0.19-0.28 | 10 | 82.2 | 0.24 | 0.21-0.28 | 4 | 53.8 | 77.50 |
| Father-offspring | 0.20 | 0.16-0.23 | 5 | 60.8 | 0.25 | 0.23-0.28 | 4 | 0.00 | 68.10 |

SMD, standardized mean difference; CI, confidence interval

$I^2$ = proportion of total variation in effect estimate due to between-study heterogeneity (based on Q)

**Table S13. Subgroup analyses by maternal BMI measurement time point**

| Family relationship | Before-pregnancy | | | | Post-pregnancy | | | | Overall $I^2$ (%) |
|---|---|---|---|---|---|---|---|---|---|
| | SMD | 95%CI | Study number | $I^2$ (%) | SMD | 95%CI | Study number | $I^2$ (%) | |
| Mother-daughter | 0.27 | 0.23-0.31 | 4 | 0.00 | 0.24 | 0.19-0.29 | 6 | 80.5 | 70.5 |
| Mother-son | 0.21 | 0.15-0.27 | 5 | 40.8 | 0.24 | 0.20-0.28 | 6 | 70.0 | 61.8 |
| Mother-offspring | 0.25 | 0.20-0.29 | 6 | 39.3 | 0.23 | 0.19-0.27 | 8 | 85.9 | 76.6 |

SMD, standardized mean difference; CI, confidence interval

$I^2$ = proportion of total variation in effect estimate due to between-study heterogeneity (based on Q)

**Table S14. Subgroup analyses by offspring age**

| Family relationship | Early adulthood | | | | Mid-adulthood | | | | Late-adulthood | | | | Overall I² (%) |
|---|---|---|---|---|---|---|---|---|---|---|---|---|---|
| | SMD | 95%CI | Study number | I² (%) | SMD | 95%CI | Study number | I² (%) | SMD | 95%CI | Study number | I² (%) | |
| Mother-daughter | 0.24 | 0.20-0.28 | 7 | 51.7 | NA | NA | NA | NA | 0.26 | 0.18-0.34 | 3 | 87.2 | 70.5% |
| Mother-son | 0.23 | 0.19-0.26 | 8 | 41.0 | NA | NA | NA | NA | 0.24 | 0.16-0.31 | 3 | 85.4 | 61.8% |
| Father-daughter | 0.25 | 0.21-0.30 | 4 | 16.3 | NA | NA | NA | NA | 0.16 | 0.11-0.21 | 3 | 51.8 | 68.5% |
| Father-son | 0.26 | 0.23-0.29 | 4 | 0.00 | NA | NA | NA | NA | 0.25 | 0.18-0.31 | 3 | 72.5 | 39.9% |
| Mother-offspring | 0.22 | 0.19-0.26 | 9 | 66.9 | 0.27 | 0.23-0.32 | 2 | 0.00 | 0.25 | 0.17-0.33 | 3 | 93.2 | 83.3% |
| Father-offspring | 0.24 | 0.19-0.28 | 5 | 57.6 | 0.22 | 0.16-0.28 | 1 | NA | 0.21 | 0.16-0.26 | 3 | 76.4 | 40.2% |

*Early adulthood: 18-30y; Mid-adulthood: 25-39y or 30-40y; Late adulthood: >40y

SMD, standardized mean difference; CI, confidence interval; NA, not available

$I^2$ = proportion of total variation in effect estimate due to between-study heterogeneity (based on Q)

**Table S15. Sensitivity analyses-standardized mean difference***

| Series | Family relationship | SMD | 95%CI | I² (%) | Study number |
|---|---|---|---|---|---|
| **Level1** | Mother-daughter | 0.25 | 0.20-0.30 | 81.8 | 10 |
| | Mother-son | 0.24 | 0.20-0.28 | 61.6 | 14 |
| | Father-daughter | 0.19 | 0.15-0.23 | 63.1 | 10 |
| | Father-son | 0.22 | 0.17-0.26 | 64.6 | 12 |
| **Level2** | Mother-offspring | 0.25 | 0.21-0.29 | 86.2 | 16 |
| | Father-offspring | 0.21 | 0.17-0.26 | 91.6 | 13 |

*unadjusted SMD, exclude studies with some participants younger than 18 years.

SMD, standardized mean difference; CI, confidence interval

$I^2$ = proportion of total variation in effect estimate due to between-study heterogeneity (based on Q)

**Table S16. Sensitivity analysis- standardized mean difference taking out studies with low quality score***

| Series | Family relationship | Pooled SMD for adjusted models | | | | Pooled SMD for unadjusted models** | | | |
|---|---|---|---|---|---|---|---|---|---|
| | | SMD | 95%CI | $I^2$ (%) | Study number | SMD | 95%CI | $I^2$ (%) | Study number |
| **Level1** | Mother-daughter | 0.25 | 0.21-0.29 | 68.6 | 10 | 0.26 | 0.21-0.31 | 82.6 | 15 |
| | Mother-son | 0.23 | 0.20-0.26 | 61.8 | 11 | 0.24 | 0.21-0.27 | 59.6 | 16 |
| | Father-daughter | 0.21 | 0.16-0.26 | 67.9 | 7 | 0.20 | 0.16-0.25 | 70.0 | 11 |
| | Father-son | 0.25 | 0.21-0.28 | 38.5 | 7 | 0.22 | 0.18-0.26 | 82.6 | 13 |
| **Level2** | Mother-offspring | 0.24 | 0.22-0.27 | 74.7 | 12 | 0.25 | 0.22-0.29 | 85.5 | 17 |
| | Father-offspring | 0.23 | 0.20-0.26 | 65.0 | 8 | 0.22 | 0.18-0.25 | 73.7 | 12 |

SMD, standardized mean difference; SD, standardized deviation; CI, Confidence interval

*take out studies by Kvaavik et al.[13], Cho et al.[35], and Khoury et al.[1]

**include studies reporting correlation and unadjusted SMD in regression coefficient

$I^2$ = proportion of total variation in effect estimate due to between-study heterogeneity (based on Q)

**Table S17. Study quality assessment using Adapted Newcastle-Ottawa scale**

| Study, year | Selection | | | Comparability | Outcome | | | Stars |
|---|---|---|---|---|---|---|---|---|
| | Representative-ness of the exposed cohort | Selection of the non-exposed cohort | Ascertainment of exposure | Comparability of cohorts on the basis of the design or analysis | Assessment of outcome | Was follow-up long enough for outcomes to occur | Adequacy of follow up of cohorts | |
| Khoury et al. [1] 1983,USA | --- | ★ | --- | --- | --- | ★ | --- | 2 |
| Friedlanderet al.[2] 1988, Israel | ★ | ★ | ★ | ★ | ★ | ★ | --- | 6 |
| Rotimi et al.[4] 1995,USA | ★ | ★ | ★ | ★ | ★ | ★ | --- | 6 |
| Lake et al.[5] 1997,UK | ★ | ★ | ★ | ★ | ★ | ★ | ★ | 7 |
| Al-Isa et al.[6] 1998, Kuwaiti | --- | ★ | --- | ★ | ★ | ★ | ★ | 5 |
| Burke et al.[7] 2001, Australia | --- | ★ | ★ | ★ | ★ | ★ | --- | 5 |
| Williams et al.[8] 2001,New Zealand | ★ | ★ | --- | ★ | ★ | ★ | ★ | 6 |

| | | | | | | | | |
|---|---|---|---|---|---|---|---|---|
| Laitinen et a[9] 2001,Finland | ★ | ★ | ★ | ★ | ★ | ★ | ★ | 7 |
| Magnusson et al.[10] 2002,Sweden | ★ | ★ | --- | ★ | --- | ★ | ★ | 5 |
| Salces et al.[11] 2002, Spain | --- | ★ | ★ | --- | ★ | ★ | ★ | 5 |
| Mirmiran et al.[12] 2002, Iran | ★ | ★ | ★ | --- | ★ | ★ | --- | 5 |
| Kvaavik et al.[13] 2003, Norway | --- | ★ | --- | --- | --- | ★ | --- | 2 |
| Wu et al.[14] 2003, Taiwan | --- | ★ | ★ | ★ | ★ | ★ | --- | 5 |
| Kazumi et al.[16] 2005, Japan | --- | ★ | --- | --- | ★ | ★ | ★ | 4 |
| Crossman et al.[17] 2006, USA | ★ | ★ | --- | ★ | ★ | ★ | --- | 5 |
| Kivimäki et al.[18] 2007, Finland | ★ | ★ | ★ | ★ | ★ | ★ | --- | 6 |
| Abu-Rmeileh, Hart et al. [19] 2008, Scotland | ★ | ★ | ★ | ★ | ★ | ★ | ★ | 7 |

| | | | | | | | | |
|---|---|---|---|---|---|---|---|---|
| Koupil et al.[20], 2008, Sweden | ★ | ★ | ★ | ★ | ★ | ★ | ★ | 7 |
| Tequeanes et al.[21] 2009,Brazil | ★ | ★ | ★ | ★ | ★ | ★ | --- | 6 |
| Kowaleski-Jones et al.[22] 2009,USA | ★ | ★ | --- | ★ | --- | ★ | --- | 4 |
| Classen et al.[24] 2010, USA | ★ | ★ | ★ | --- | ★ | ★ | --- | 5 |
| Cooper et al.[25] 2010, UK | ★ | ★ | --- | ★ | ★ | ★ | --- | 5 |
| Reynolds et al.[26] 2010, UK | --- | ★ | ★ | ★ | ★ | ★ | --- | 5 |
| Al-Isa et al.[27] 2011,Kuwaiti | --- | ★ | --- | ★ | ★ | ★ | --- | 4 |
| Hochner et al. [28] 2012, Israel | ★ | ★ | ★ | ★ | ★ | ★ | --- | 6 |
| Johnson et al.[29] 2012,UK | ★ | ★ | ★ | ★ | ★ | ★ | --- | 6 |
| Murrin et al.[30] 2012, Ireland* | ★ | ★ | --- | --- | --- | ★ | --- | 3 |

| | | | | | | | | |
|---|---|---|---|---|---|---|---|---|
| Wang et al.[31] 2012, China | ★ | ★ | --- | ★ | ★ | ★ | ★ | 6 |
| Hu et al.[32] 2013, China | ★ | ★ | ★ | ★ | ★ | ★ | --- | 6 |
| Magarey et al.[15] 2003,Australia | ★ | ★ | ★ | ★ | ★ | ★ | --- | 6 |
| Kelly et al.[33] 2014,Ireland* | ★ | ★ | ★ | --- | --- | ★ | --- | 4 |
| Cho et al.[35] 2015,USA | --- | ★ | --- | --- | --- | --- | --- | 1 |
| Derraik et al. [36] 2015,Sweden | ★ | ★ | ★ | ★ | ★ | ★ | --- | 6 |
| Han et al[37] 2015,Scottland | ★ | ★ | ★ | ★ | ★ | ★ | --- | 6 |
| Alati et al.[39] 2016,Australia | ★ | ★ | ★ | ★ | ★ | ★ | --- | 6 |
| Zalbahar et al.[40] 2016,Australia | ★ | ★ | --- | ★ | ★ | ★ | ★ | 6 |
| Rath et al.[41] 2016,Australia | ★ | ★ | --- | ★ | ★ | ★ | --- | 5 |
| Swanton et al.[42] 2017,USA | ★ | ★ | ★ | ★ | ★ | ★ | --- | 6 |

| | | | | | | | | |
|---|---|---|---|---|---|---|---|---|
| Rodrigo M. Carrillo-Larco et al.[43]<br>2018,Peru | ★ | ★ | ★ | ★ | ★ | ★ | --- | 6 |
| Kaseva et al.[44]<br>2018,Finland | --- | ★ | ★ | --- | ★ | ★ | --- | 4 |
| Schoppa et al.[45]<br>2018,Finland | ★ | ★ | ★ | ★ | ★ | ★ | --- | 6 |
