## Appendix for "Body mass index in parents and their adult offspring: a systematic review and meta-analysis"

Search strategies

Adapted Newcastle–Ottawa scale for cohort studies

**Appendix**

Search strategies

| Database | Search Query | Results |
| --- | --- | --- |
| PubMed | #1,"Search ((((((((body mass index[MeSH Terms]) OR body weight[MeSH Terms]) OR body size[MeSH Terms]) OR body composition[MeSH Terms]) OR body constitution[MeSH Terms]) OR adiposity[MeSH Terms]) OR overweight[MeSH Terms]) OR obesity[MeSH Terms]) OR bmi[Title/Abstract] Sort by: [pubsolr12]" | 717,217 |
|  | #2,"Search ((((((offspring[Title/Abstract]) OR adult child[Title/Abstract]) OR adult children) OR adolescent) OR adult daughter) OR adult son) OR young adult Sort by: [pubsolr12]" | 2,634,623 |
|  | #3 ,"Search (((((((((parental[Title/Abstract]) OR maternal[Title/Abstract]) OR prepregnancy[Title/Abstract]) OR pre-pregnancy[Title/Abstract]) OR father[Title/Abstract]) OR mother[Title/Abstract]) OR parents[Title/Abstract]) OR male[Title/Abstract]) OR female[Title/Abstract]) OR paternal[Title/Abstract] Sort by: [pubsolr12]" | 1,736,959 |
|  | #4,"Search (#2) AND #3 Sort by: [pubsolr12]" | 145,586 |
|  | #5,"Search (#1) AND #4 Sort by: [pubsolr12]" | 137,988 |
|  | #6,"Search (#5) AND #6 Sort by: [relevance]", | 39,253 |
|  | #7,"Search (#5) AND #6 Filters: Humans Sort by: [pubsolr12]", | 32,689 |
| EmBase | #1 ('body mass index':ab,ti OR 'body weight':ab,ti OR 'body size':ab,ti OR 'body composition':ab,ti OR 'body constitution':ab,ti OR adipsoity:ab,ti OR obesity:ab,ti OR overweight:ab,ti OR bmi:ab,ti) AND [1980-2020]/py | 980,121 |
|  | #2 (offspring:ab,ti OR 'adult child':ab,ti OR 'adult son':ab,ti OR 'adult daughter':ab,ti OR 'young adult':ab,ti OR 'old adult':ab,ti OR 'adolescents'/exp OR adolescents) AND [1980-2020]/py | 383,832 |
|  | #3 (parental:ab,ti OR maternal:ab,ti OR paternal:ab,ti OR parents:ab,ti OR father:ab,ti OR mother:ab,ti OR male:ab,ti OR female:ab,ti OR prepregnancy:ab,ti) AND [1980-2020]/py | 2,6664,995 |
|  | #4 #1 AND #2 | 16,410 |
|  | #5 #1 AND #3 | 236,677 |
|  | #6 #4 AND #5  Total | 12,006  32,689 |

### Adapted Newcastle–Ottawa scale for cohort studies ^1^

**Selection**

1) Representativeness of the exposed cohort ^1^

a) truly representative of the average maternal OR paternal population^a^ in the community **🟑**

b) somewhat representative of the average maternal OR paternal population^a^ in the community **🟑**

c) selected group of users eg ^a^

d) no description of the derivation of the cohort

2) Selection of the non-exposed cohort ^1^

a) drawn from the same community as the exposed cohort **🟑**

b) drawn from a different source

c) no description of the derivation of the non exposed cohort

3) Ascertainment of exposure ^2^

a) secure record (eg explicitly measured weight and height^a^) **🟑**

b) structured interview e.g. validated self-report^a^**🟑**

c) any self-report

d) no description

**Comparability**

4) Comparability of cohorts on the basis of the design or analysis ^3^

a) study controls for gestational diabetes^a^ **🟑**

b) study controls for any additional factor **🟑**

c) no factors controlled for^a^

**Outcome** (note: outcome is child weight status)

5) Assessment of outcome ^2^

a) independent blind assessment **🟑**

b) record linkage **🟑**

c) self-report

d) no description

6) Was follow-up long enough for outcomes to occur ^2^

a) yes (select an adequate follow up period for outcome of interest: all children in the analysis were the target age of the research question^a^) **🟑**

b) no

7) Adequacy of follow up of cohorts ^1^

a) complete follow up - all subjects accounted for **🟑**

b) subjects lost to follow up unlikely to introduce bias - small number lost – >80%^a^ follow up, or description provided of those lost) **🟑**

c) follow up rate < 80%^a^ and no description of those lost

d) no statement

Total number of stars (out of a possible 8 stars^b^):

Note: A study can be awarded a maximum of one star for each numbered item within the Selection and Outcome categories. A maximum of two stars can be given for Comparability.

Footnote:

^a^Red font is where form was adapted to make questions relevant to this systematic review.

^b^Original question 4 “Demonstration that outcome of interest was not present at start of study” in the original scale is not applicable to child weight status outcomes as women are identified in early pregnancy using their pre/early pregnancy BMI and their offspring weight data do not exist at the start of the study. Therefore, this item has been removed from the scale. The denominator value for the maximum number of stars a study can be awarded has been reduced from 9 to 8 due to the removal of the original question 4.

^1^Questions assessing selection bias

^2^Questions assessing information bias

^3^Questions assessing confounding
